# Frequent introductions and climate suitability drive increasing dengue risk in Florida

**DOI:** 10.64898/2026.05.01.26352185

**Authors:** Emma Taylor-Salmon, Yi Ting Chew, Rafael Lopes, Timothy Locksmith, Edgar Kopp, Julieta Vergara, Amanda Davis, Molly Mitchell, Pamela Colarusso, Sarah Schmedes, Valerie Mock, Blake Scott, Rebecca Zimler, Chalmers Vasquez, Maday Moreno, Lauren M. Paul, Scott F. Michael, Mallery I. Breban, Chantal B.F. Vogels, Joshua L. Warren, Colin J. Carlson, Danielle Stanek, Lea Heberlein, Verity Hill, Andrea Morrison, Nathan D. Grubaugh

## Abstract

In recent years, detection of local dengue cases in Florida have increased in both frequency and geographical extent. From 2022 to 2024, consecutive outbreaks in Miami-Dade County were mainly caused by a single lineage of dengue virus (DENV) serotype 3, prompting questions about changing epidemiology and a transition towards endemicity. In this study, we used mathematical modeling and genomic epidemiology to reveal the spatiotemporal dynamics and drivers of local dengue cases in Florida. We found that annual clusters and outbreaks were caused by frequent short-lived DENV introductions, primarily from the Caribbean, and did not find evidence for local trans-seasonal DENV lineage persistence. Further, we show that the climate-driven increases in local suitability for *Aedes aegypti* transmission and travel-associated cases were the greatest risk factors for outbreaks in Miami-Dade and the geographic expansion of dengue in Florida. Overall, while we do not yet find evidence for endemicity, we demonstrate how climatic trends are enhancing the local public health risk caused by dengue in Florida.

## Introduction

Dengue is an acute febrile illness caused by four genetically- and antigenically-related, but distinct, dengue virus (DENV) serotypes (*1*). DENV is transmitted to humans by *Aedes* mosquitoes: primarily by *Ae. aegypti,* with *Ae. albopictus* mostly as a secondary vector (*2*). Dengue is the most common arboviral disease worldwide; half of the world resides in endemic areas, with children and adolescents bearing a majority of the disease burden (*3*, *4*). In 2024, PAHO reported the highest dengue incidence recorded in the Americas, with almost 7 million confirmed and more than 13 million suspected dengue cases (*5*). This easily surpassed 2023, the previous high at 2 million confirmed and 4.5 million suspected cases (*5*). This increase of dengue in the region is also leading to increases in travel-associated and subsequent local dengue incidence in the contiguous United States (US), with 366 locally acquired cases reported from 2022-2024 (*6*). Over 90% of the recent surge in dengue has occurred in Florida, where not only has Miami-Dade County experienced larger outbreaks, but the number of Florida counties reporting local dengue cases has also expanded.

The Florida Department of Health (FDOH) has a robust dengue surveillance system, which captures symptomatic patients with local or travel-associated infections (*7*). Since 2009, Florida has reported an increasing number of travel-associated cases (*8*). Despite reporting no locally acquired dengue cases from 1934 to 2009, the FDOH has detected numerous local outbreaks over the past two decades (*7*). While it is known that infected travelers are important sources of local DENV transmission in Florida (*9–11*), it is not clear if, or how, the interplay between local climate suitability for mosquitoes and virus introductions is changing. Specifically, there is an urgent need to understand what currently drives dengue outbreaks, and whether dengue is transitioning towards endemicity in Florida.

In this study, we combined case surveillance, virus sequencing, and transmission dynamic modeling to estimate the dynamics and drivers of DENV in Florida from 2009 to 2024. First, we show that local dengue incidence has been increasing since 2009 and expanding from southern into central Florida. Then, using sequencing data, we performed phylogeographic analyses to determine the sources and persistence of local DENV lineages. Finally, we constructed spatiotemporal models to understand why local dengue cases primarily occur in southern Florida counties and what drives the transmission dynamics in Miami-Dade. Overall, our study highlights the importance of local dengue surveillance and genomic epidemiology in understanding DENV transmission and spread within Florida.

## Results

### Recent expansion of local dengue cases in Florida

Across the Americas, there has been a recent surge of dengue cases (*5*). Likewise, the US has experienced a rapid increase in the number of travel-associated dengue cases, with Florida reporting 3,713 travel-associated cases from 2010-2024 (*7*) (**Fig. 1A**). In 2009, the first locally-acquired dengue case in Florida since 1934 was reported in Monroe County (*12*, *13*), with a total of 22 reported cases that year (**Fig. 1B**). The outbreak stretched into 2010 (*12–14*), with another 66 local cases diagnosed, as well as two additional cases in Broward (1) and Miami-Dade (1) counties. This was followed by several years with scattered local cases reported mostly in the southern portion of the state **(Fig. S1**). However, in 2013 there was another local dengue outbreak, this time in Martin County with 24 reported cases (*10*), as well as 2 other cases reported in Miami-Dade County (**Fig. 1B**). Afterwards, there were once again minimal local dengue cases reported from 2014-2018 (**Fig. S1**). Yet, as dengue cases rebounded throughout the Americas following the Zika epidemic (*15*), so did local cases within Florida, with larger outbreaks and more counties involved. In 2020, Monroe County experienced its largest reported outbreak (72 cases) and its first in a decade (**Fig. 1B**), but Miami-Dade County was the focal point of recent dengue activity (*11*). The first local dengue case in Maimi-Dade County was reported in 2010 but the county did not exceed ten reported cases within a year until 2019 (16 cases; **Fig. 1B, Fig. S1**). After that, local dengue cases reported from Maimi-Dade County started to increase with 64 in 2022 (incidence rate = 2.3 per 100,000 population), 173 in 2023 (6.2 per 100,000), and 50 in 2024 (1.8 per 100,000; **Fig. 1B**). From 2009 through 2024, there were a total of 594 local dengue cases reported from 15 counties (**Fig. 1C**), with the last 3 years responsible for 360 (61%) of those cases and 8 counties reporting their first local case (**Fig. 1B**). This concerning trend highlights that the opportunities and suitable conditions for local DENV transmission in Florida are expanding.

**Figure 1.**
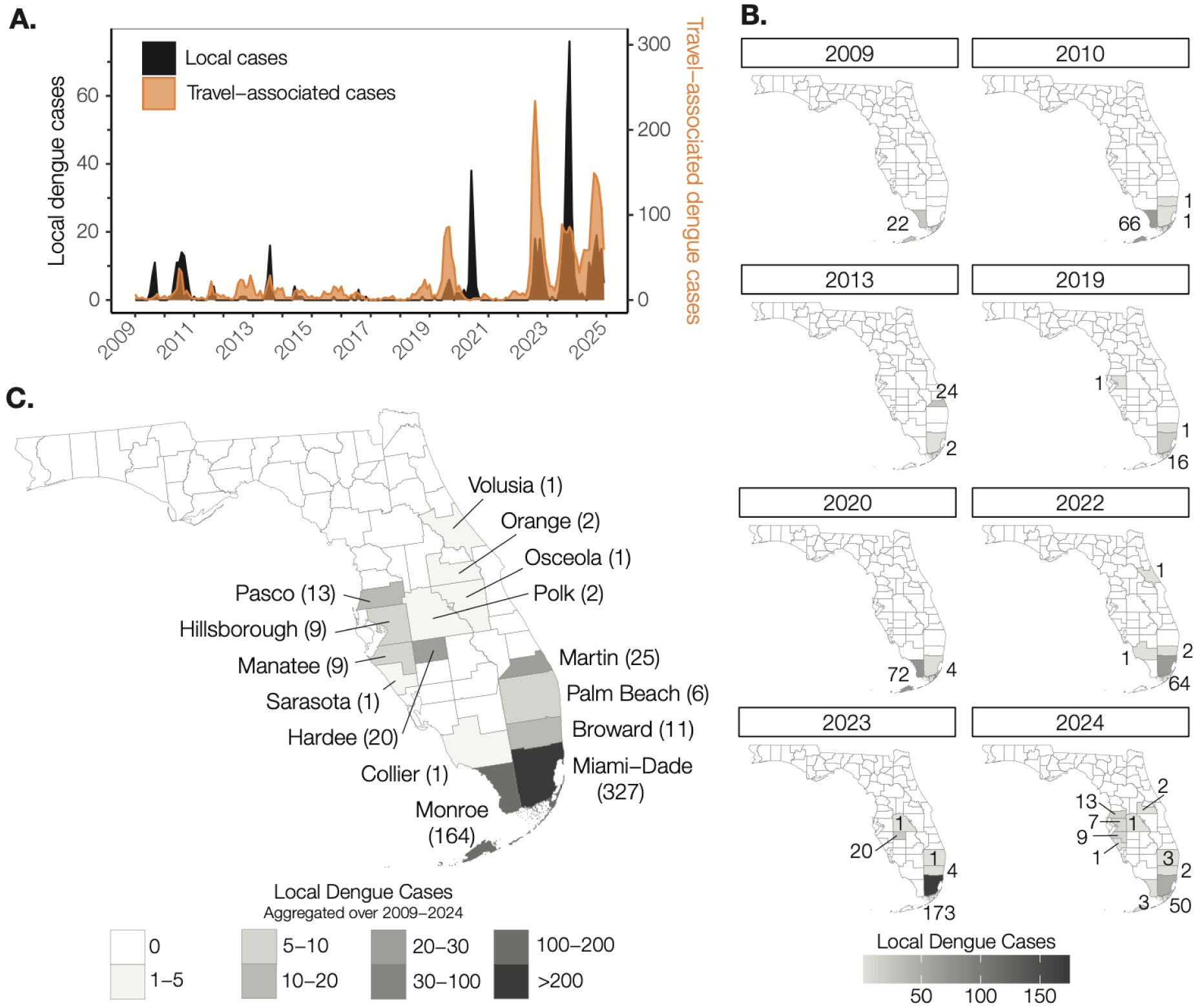
Local dengue cases have been increasing, especially in southern Florida, from 2009 to 2024. (**A**) Monthly local dengue cases and monthly travel-associated dengue cases from Florida. (**B**) Annual number of local dengue cases per county reported by FDOH. Only years with > 10 local cases were included. The complete data can be found in Supplementary Figure S1). (**C**) Aggregated local case data for each county during the study.

### Phylogenetics reveals patterns of short-lived dengue virus introductions and no trans-seasonal persistence

To reconstruct the transmission dynamics behind local dengue outbreaks in Florida, we sequenced 133 local and 294 travel dengue cases from June 2010 through September 2024 (**Table S2**) and conducted discrete phylogeographic analyses (global DENV genomic dataset n=5,580, **Table S1**). This type of phylogenetic analysis allowed us to estimate the times and sources of DENV introductions and the duration of local persistence within Florida (*16*, *17*). We conducted one analysis for each DENV serotype, grouping locations with varying spatial resolutions to highlight transmission dynamics between counties in Florida, regions in the Americas, and elsewhere (**Table S1**). Consistent with earlier studies (*9*), our data revealed that DENV dynamics in Florida are driven by frequent introductions that sometimes lead to secondary local transmission, but not by endemic trans-seasonal persistence (**Fig. 2**).

**Figure 2.**
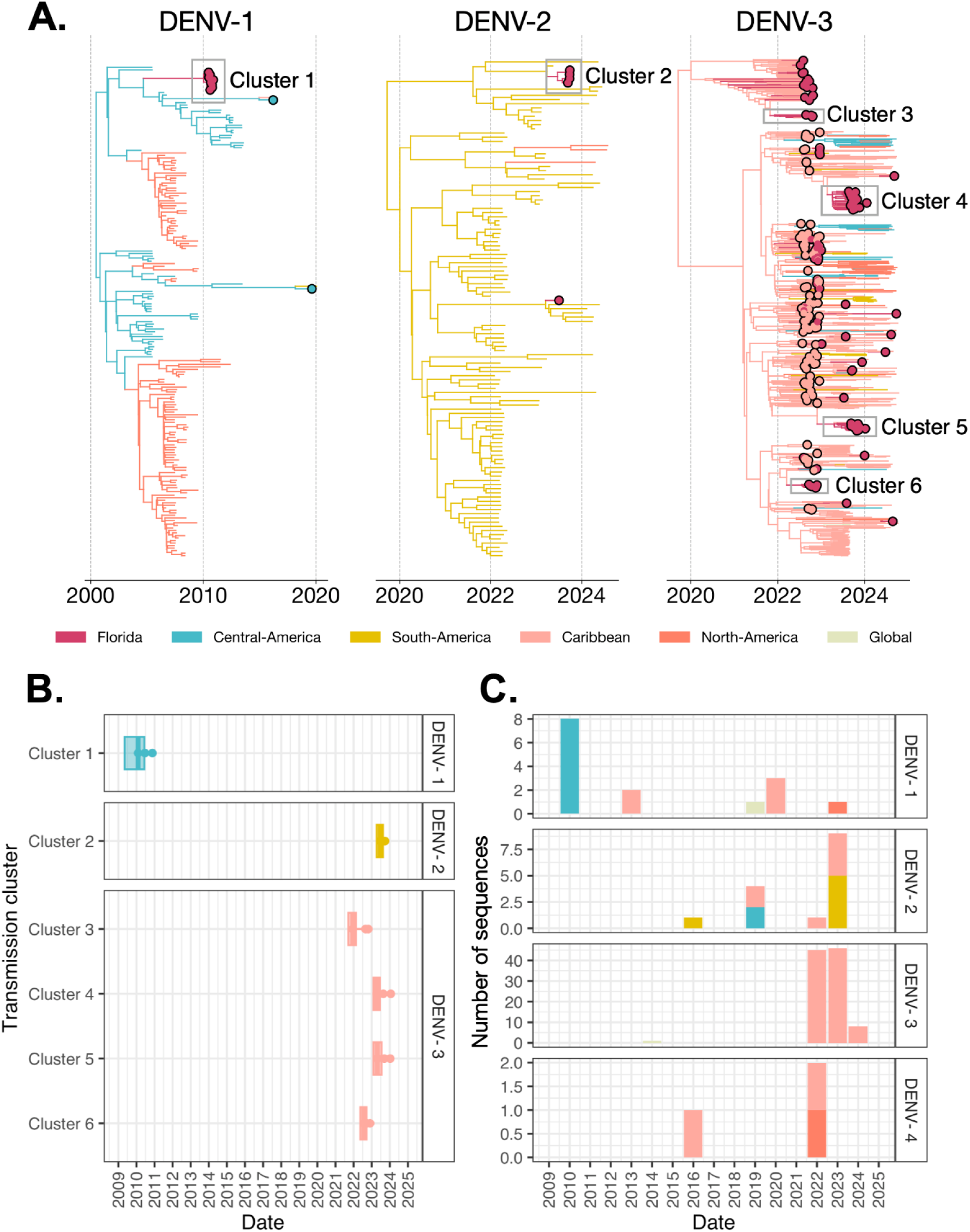
Local dengue cases in Florida are due to short-lived lineage introductions, not underlying endemic transmission. (**A**) Time-resolved phylogenies of each serotype, with the branches and tips colored by inferred source and observed sampled regions, respectively. Larger circles represent DENV samples from locally-acquired and travel-associated cases in Florida. Transmission clusters of locally-acquired samples are outlined in black boxes. The phylogenetic trees are shown for DENV-1, DENV-2, and DENV-3 from left to right. DENV-4 was not shown as no transmission clusters were detected, only single sequences. (**B**) Introduction time of transmission clusters (i.e., three or more locally-acquired sequences) by serotype, colored by inferred source regions. The times of the most recent common ancestor of the clusters are taken conservatively as introduction times, with 95% CIs indicated by box boundaries. (**C**) Number of sequences for each serotype for all local cases by year, colored by inferred source regions.

All four DENV serotypes were detected amongst sequenced local DENV cases, and each serotype was dominated by one major lineage: DENV-1V_C (53.3%, n=8), DENV-2II_F (60.0%, n=9), DENV-3III_B (99.0%, n=99) and DENV-4II_B (100.0%, n=3; **Table S2**). Most sequenced local cases from Florida were DENV-3III_B (74.4%, n=99), which was first sampled in Miami-Dade in July 2022 (*8*, *11*). This lineage, specifically the emerging sublineage DENV-3III_B.3.2 (*8*, *18–20*), was subsequently responsible for the consecutive DENV-3 outbreaks in Miami-Dade from 2022 to 2024 (**Fig. 1**, **Fig. 2C**). However, we did not detect local trans-seasonal (> 1 year) persistence of this lineage, and instead identified four transmission clusters (**Methods**) originating from independent introductions from the Caribbean (likely primarily from Cuba (*8*); Clusters 3-6; **Fig. 2A, 2B, Table S3**). We estimated that the earliest detected cluster of DENV-3III_B.3.2 (Cluster 3; n=4 local DENV sequences) was introduced by late 2021 to early 2022 (95% highest posterior density credible interval [CI]: September 2021 to February 2022, **Fig. 2B**), corresponding to the start of an outbreak in Cuba that spread throughout the Americas (*8*, *11*, *20*, *21*). From 2022 onwards, we detected multiple independent 3III_B.3.2 clusters of local transmission in short succession with median introduction times in July 2022 (Cluster 6, 95% CI: May to September 2022) and April 2023 (Cluster 4, 95% CI: February to June 2023; Cluster 5, 95% CI: February to July 2023; **Fig. 2B, Table S3**). Given the high number of introductions of this lineage from infected travelers in 2022 (*8*), it is not surprising that this would lead to multiple independent clusters of local transmission.

Similarly, we did not detect trans-seasonal persistence for other serotypes, but instead identified only short-lived transmission clusters. We estimated that Cluster 1 (DENV-1V_C) was introduced into Florida from Central America by late 2009 to early 2010 (95% CI: May 2009 to June 2010, **Fig. 2B**) and led to the 2010 outbreak in Monroe County. This is consistent with previous studies that analyzed viral envelope glycoprotein coding sequences from several patient samples, as well as a mosquito pool, in Monroe County in 2010 and found that it was closely related to DENV-1 strains from Nicaragua and Mexico (*14*, *22*, *23*). We estimated that Cluster 2 (DENV-2II_F) was introduced into Florida from South America by early to mid 2023 (95% CI: May to August 2023, **Fig. 2B**) and contained local Miami-Dade County cases reported in September of that year (**Table S3, Table S4**). Therefore, in 2023 there were not only multiple introductions of DENV-3III_B into Miami-Dade, there were introductions of multiple serotypes, highlighting the importance of this county when investigating DENV dynamics within Florida. We only sequenced three DENV-4 genomes from the available clinical sample set and none were phylogenetically related.

We detected introductions of all four serotypes from the Caribbean, which was the most commonly inferred introduction source and only region inferred as a source for all detected serotypes (n=113, 84.96%, **Fig. 2C, Table S4**). This is consistent with the surveillance data, as the majority of travel-associated dengue cases in Florida reported recent travel to the Caribbean (*8*). Despite more frequent local dengue cases in Florida being observed from 2022 to 2024, our analysis supports previous findings that this was not caused by DENV trans-seasonal persistence of DENV within Florida and that most local cases are instead due to recent introductions from the Americas, usually the Caribbean.

### Viral introductions and climate-driven transmission suitability govern risks of local dengue cases in Florida

We demonstrated that local dengue cases in Florida are caused by short-lived virus introductions (**Fig. 2**) despite the more widespread occurrence of local dengue cases in recent years (**Fig. 1**). To obtain a better understanding of what factors govern where local dengue cases are detected and the recent spatial expansion within Florida, we investigated the relative influence of various travel-associated (i.e., virus introductions), climatic (i.e., temperature and hydrometeorological conditions), and socioeconomic risk factors on the annual occurrence of local dengue cases at the county level (**Table S5**). Using these covariates, we constructed a spatiotemporal model of the annual occurrence of local dengue cases in Florida counties from 2009 to 2024 and found that it is mostly associated with the detection of travel cases and local climatic suitability (**Fig. 3**).

**Figure 3.**
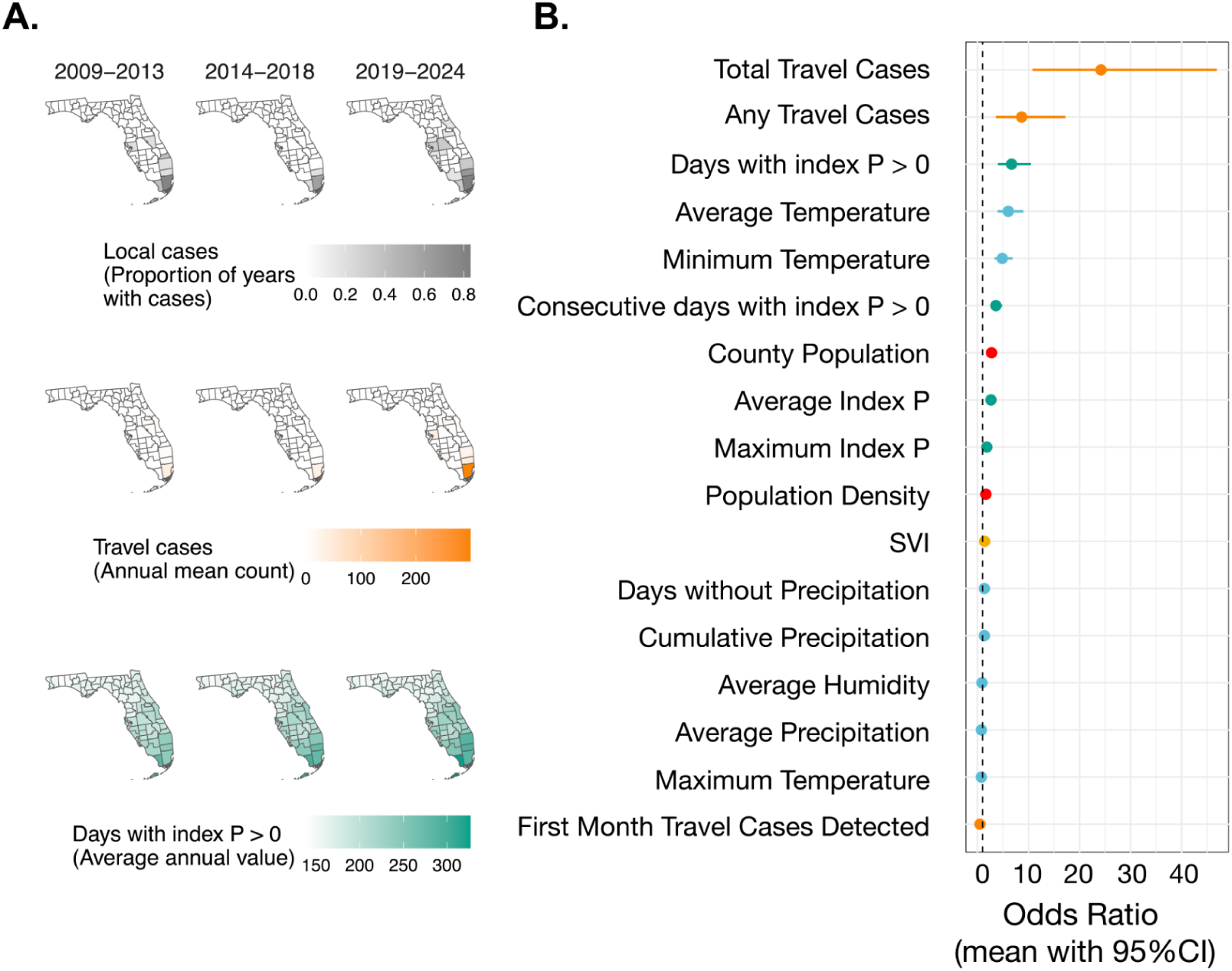
The presence of infected travelers and the yearly mean transmission suitability are the most impactful drivers of county-level local dengue occurrence in Florida. (**A**) Posterior mean and 95% credible interval for all odds ratio computed from linear fixed effects were used to describe how annual local dengue case occurrence by county is predicted by changing a covariate (e.g., mean index P). The circles indicate the posterior mean and the lines indicate the 95% credible interval. Covariates are colored by category. (**B**) Trends in spatial distribution of local dengue cases, travel cases, and index P between counties in Florida are represented by how the proportion of years with local dengue case occurrence, average annual count of travel dengue cases, and average annual mean index P values have changed within each 5-6 year time period between 2009 and 2024.

Previous studies have demonstrated that DENV transmission potential can be estimated using a mosquito-borne transmission suitability measure, referred to as index P, which is calculated by incorporating local climate data (e.g., temperature and humidity) and additional mosquito factors (e.g., lifespan, biting rate, and virus extrinsic incubation period) into a Bayesian framework (*24–27*). Thus, we used index P to estimate suitability of local climatic conditions for DENV transmission between Florida counties. We calculated daily estimated DENV transmission potential by *Ae. aegypti* mosquitoes during our study period, as well as the monthly and yearly minimum, mean and maximum index P values, for each county. Yearly mean index P demonstrated a rising trend in the southern part of the state, indicating that in principle climate conditions have become increasingly favorable for local DENV transmission (**Fig. S2**).

To investigate the potential drivers of DENV spatial expansion in Florida (**Fig. 3A**), we fitted a logistic regression model with spatial random effects to estimate the probability of annual occurrence of local dengue cases at the county level (**Fig. 3B**). To account for unexplained spatiotemporal variation in annual dengue occurrence between counties in Florida (e.g., from unmeasured factors such as sampling biases), we fit a baseline model using spatially structured and unstructured random effects specified as a Besag-York-Mollié (BYM) model (**Methods**) (*28*). To this baseline model, we added each covariate individually as a linear fixed effect and visualized their effects on county-level annual dengue occurrence using posterior means as the point estimates and 95% quantile-based CIs of their odds ratios to quantify uncertainty (**Fig. 3B**).

We found that the annual county-level occurrence of local dengue cases had strong positive associations with the annual total number of travel-associated dengue cases (estimate: 24.21, 95% CI: 10.79 - 46.90), the presence of any travel-associated dengue cases during that year (estimate: 8.66, 95% CI: 3.60 - 17.21), the annual number of days with index P exceeding 0 (estimate: 6.68, 95% CI: 4.01 - 10.47), and the annual average temperature (estimate: 6.05, 95% CI: 3.92 - 9.00; **Fig. 3B**). Annual minimum temperature, consecutive number of days with index P exceeding 0, annual county population size, annual average index P, annual maximum index P, and annual county population density had weaker positive effects on the occurrence of annual local cases. The first month in which a travel-associated case was detected had a weak negative effect on annual local dengue case occurrence.

Covariates that did not have a significant impact on the annual occurrence of local dengue cases in Florida at the county-level included social vulnerability index (SVI), annual maximum temperature, or any precipitation-related variables (e.g., days without precipitation, average precipitation, cumulative precipitation).

The majority of local dengue cases from our study period were reported in Miami-Dade County (55.4%) and from the final two years (2023 and 2024, 49%). To understand how much, if any, these variables could skew our findings, we performed two sensitivity analyses by removing any cases from the years 2023 and 2024 (**Fig. S3A**) or from Miami-Dade County (**Fig. S3B**). When the years 2023 and 2024 were excluded from the analysis, we found similar associations to the initial model (**Fig. S3A**). When Miami-Dade County was excluded from the analysis, the one notable difference was that the total number of travel-associated dengue cases now has a relatively smaller positive effect on annual local dengue case occurrence (estimate: 16.30, 95% CI: 6.84 - 33.26; **Fig. S3B**).

Our model of local dengue case occurrence provides a clearer picture of the drivers of local cases within Florida, highlighting the importance of DENV introductions from infected travelers combined with annual conditions suitable for mosquito transmission.

### Travel cases drive local dengue risk in Miami-Dade with increasing climatic suitability

We demonstrated the presence of DENV-infected travelers and suitable DENV transmission potential for *Ae. aegypti* primarily governed the spatial structure of local dengue cases in Florida (**Fig. 3**). While 15 counties have reported local dengue cases, however, fewer than half have reported cases in multiple years (*n*=7). Miami-Dade County is an outlier, having reported local dengue cases from 12 of the 16 years and accounting for more than half of the cases during our study period (*n*=329, 55%). It is also the only county in which we detected multiple clusters of local transmission, both DENV-2II_F (cluster 2) and DENV-3III_B (clusters 3-6, **Fig. 2A & 2B**), which were estimated to have multiple introductions spanning from 2021 to 2023 (**Table S3**).

Furthermore, Miami-Dade County has only recently experienced large local dengue outbreaks, in that most of its local cases were detected between 2022 and 2024 (n=287, 87%). Thus, separately from our occurrence model described above (**Fig. 3**), we developed an incidence model to evaluate the effects of travel cases, mosquito abundance, and climatic covariates on the monthly incidence of local dengue cases in Miami-Dade County (**Fig. 4**).

**Figure 4.**
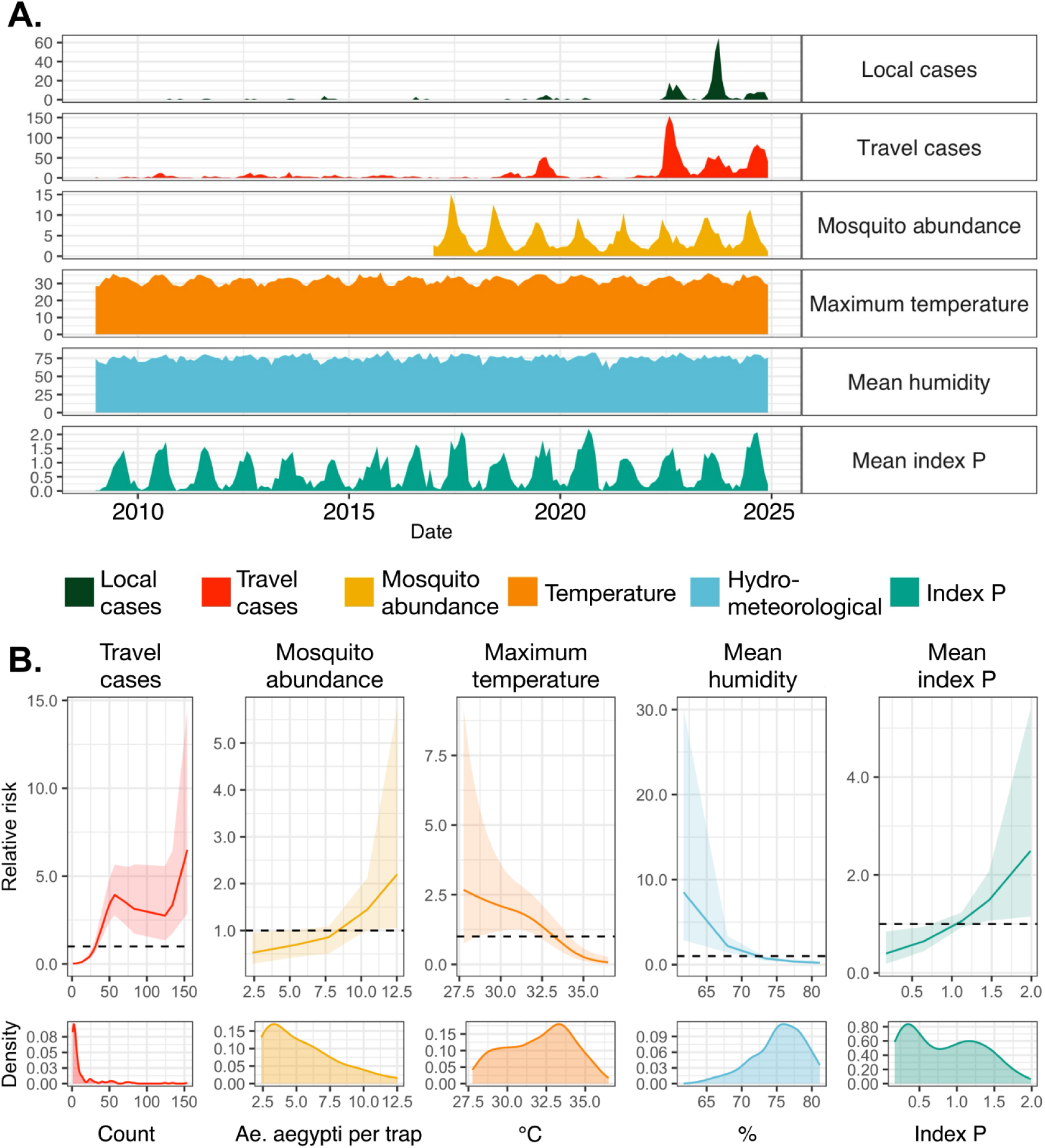
Monthly local dengue incidence in Miami-Dade from 2009 to 2024 are mostly driven by higher travel cases and climatic suitability for local virus transmission. (A) Time series of monthly local cases, modeled by covariates capturing travel cases detected, mosquito abundance, temperature, humidity, and index P values. (B) Non-linear marginal effects of all covariates from our full model are shown on the relative risk scale with their posterior mean and 95% credible interval shown as lines and ribbons respectively. Observed distributions are plotted as probability densities on the bottom-most row. All covariates used for modeling are entered at monthly resolutions, grouped broadly, and colored by type: travel-associated dengue cases (red), *Ae. aegypti* per trap (yellow), temperature (orange), hydrometeorological variables (blue), and index P (green).

Within Miami-Dade County, *Ae. aegypti* is the predominant DENV vector in Miami-Dade County (*29*) and is detected there year-round (*30*). The Miami-Dade County Mosquito Control Division has a standardized mosquito surveillance system, which allowed us to obtain monthly *Ae. aegypti* trap data in Miami-Dade County from 2017-2024, encompassing the time period during which most of the local cases were reported (n=310, 94%). We incorporated these data into a monthly incidence model of dengue cases in Miami-Dade County (**Fig. 4**). For this model, we aggregated climatic covariates from daily observed values to obtain monthly statistical values for temperature (mean, minimum, maximum), travel cases (count), precipitation (mean, total, number of days without precipitation), relative humidity (mean), and index P (mean, maximum, number of days with index P > 0; **Fig. 4A**).

We found that the monthly incidence of local dengue cases in Miami-Dade County had a positive and non-linear relationship with the total monthly travel cases, such that the relative risk of local dengue cases exceeded 1 when there were greater than ∼31 travel cases/month (**Fig. 4B**). The maximum relative risk (RR: 6.5, 95% CI: 2.9 - 14.5) occurred at 154 travel cases/month. Mosquito abundance had a weak positive relationship with monthly local dengue incidence, with relative risk exceeding 1 when there were > 10 *Ae. aegypti* per trap, and was maximized at 12.5 *Ae. aegypti* per trap (RR: 2.2, 95%CI: 1.0 - 5.7).

Temperature and humidity had weak negative relationships (**Fig. 4B**), which we modelled here without time lags. The relative risk of monthly local dengue incidence decreased below 1 for monthly maximum temperatures > 33.4°C and is maximized at 27.8°C (RR: 2.67, 95% CI: 0.8 - 9.2). Our results may reflect the delayed effects of temperature on local dengue cases, but also may be because there is an optimal range of temperature values for local dengue transmission by *Ae. aegypti* (*31–33*). Thermal response experiments and mechanistic models estimate that transmission peaks around 29.1 °C and declines to zero for temperatures below 17.8 °C or greater than 34.6 °C (*31*). Relative risk of monthly local dengue incidence decreased below 1 when monthly mean humidity was greater than 73.3%, and was maximized at 61.7% (RR: 8.5, 95% CI: 2.9 - 30.0).

Combining both local temperature and humidity data with the appropriate time lags, we found that monthly mean index P had a positive monotonic effect, with relative risk of monthly local dengue incidence exceeding 1 for monthly mean index P > 1.12 (**Fig. 4B**). Index P exhibited seasonal variation, similar to what was observed with other climatic covariates (i.e., temperature, humidity) and mosquito abundance, but was slightly more informative in its positive monotonic relationship with monthly local incidence (95% CI for RR spans 1 for mean index P values between 1.1 - 1.5).

## Discussion

### Local surveillance and genomic epidemiology reveal drivers of local dengue cases

By analyzing viral genomics and surveillance data, we uncovered important drivers of the recent expansion of local dengue cases within Florida. Dengue cases resulting from local transmission have increased in Florida over the last two decades, especially in the southern part of the state (**Fig. 1**). We show that clusters of local cases are due to new DENV introductions and not trans-seasonal persistence from prior outbreaks (**Fig. 2**). Moreover, we found that the annual occurrence of local dengue cases in Florida counties is associated with the numbers of reported infected travelers and high annual mosquito transmission potential (**Fig. 3**), highlighting the importance of both viral introductions and local climatic suitability for DENV transmission by *Ae. aegypti* when understanding local dengue risk in Florida. Finally, we estimated the impact of different travel, mosquito, and climatic covariates on monthly local dengue incidence in Miami-Dade County in which more than half of all local dengue cases in Florida were detected (**Fig. 4**) and was also the most affected area during the 2016 Zika outbreak (*34*, *35*). From this, we estimate that local dengue cases reported from Miami-Dade County are positively associated with travel-associated cases and mosquito transmission potential. Overall, we demonstrate that DENV introductions via infected travelers are the main driver of the notable increase in local dengue cases throughout Florida, meaning the regional DENV dynamics have a direct impact on local public health in Florida.

### Local transmission in Florida is increasing, but has not established endemicity

During the period of increased local transmission in Miami-Dade County from 2022-2024, we detected five distinct (one DENV-2II_F and four DENV-3III_B.3.2) phylogenetic clusters that were all short-lived, going extinct within a year. Though for clusters that started in 2024, we do not yet have data to determine if they persisted into 2025. Four of these clusters were introduced from the Caribbean, which is not surprising given the high levels of transmission (*5*) coupled with the large number of travel-associated infections in Florida that report recent travel from that region (*7*, *8*). While suitable climatic conditions for local transmission are necessary for outbreaks, the recent increase in dengue incidence in Miami-Dade County did not coincide with higher levels of mosquito abundance, transmission suitability (index P), and other DENV-relevant climatic variables (e.g. temperature, humidity) relative to preceding years (**Fig. 4A**). Rather, we show that the recent increase in travel-associated cases, mostly from the Caribbean, was the greatest risk factor for local dengue cases (**Fig. 4B**).

While we did not detect trans-seasonal persistence for any of the recent clusters, it is possible that this may have occurred for earlier outbreaks. The 2009 and 2010 dengue outbreaks in Monroe County were both caused by DENV-1 (*14*). Analysis of only envelope sequences found that the samples from both years clustered together, suggestive of trans-seasonal persistence (*23*). As there are currently no available DENV sequences from 2009 that met our inclusion criteria of 70% genome coverage, we could not directly investigate lineage persistence between the outbreaks. Instead, we estimated the introduction time for the 2010 outbreak viruses (Cluster 1) to have occurred between mid-2009 to mid-2010 (**Fig. 2B**), which spans the earliest reported dates for the first local cases in Monroe County in July 2009. Thus we cannot rule out persistence. However, there is no evidence for established endemicity of this DENV-1 lineage beyond 2010. The next most recent DENV-1 outbreak in Florida occurred in 2013 in Martin County (*10*). Again, analysis of DENV envelope sequences from 2013 Martin County cases showed that the viruses were most closely related to South American DENV-1 sequences that expanded into the Caribbean (*10*), and were not the result of persistence of DENV-1 from Monroe County a few years earlier.

Thus, we show that DENV has not yet established persistent endemic transmission in Florida despite the numerous outbreaks and more frequent opportunities. Critically, research is needed to determine if and when the conditions are suitable for endemicity as this transition would significantly increase the public health risk caused by DENV. For example, the local dengue incidence rate from Miami-Dade County from 2022-2024 ranged from 2-6 cases per 100,000 population. Under endemic transmission scenarios closer to what is experienced in Puerto Rico during the same time period, 30-190 cases per 100,000 population (*5*), the local dengue cases in Miami-Dade County (population 2.8 million) could climb into the thousands.

### Climate driven expansion of transmission suitability in Florida

The location of local dengue case reports in Florida have been expanding, with eight of the fifteen counties with local dengue cases from 2009-2024 having reported their first case from 2022-2024 (Collier, Hardee, Manatee, Orange, Pasco, Polk, Sarasota, and Volusia, **Fig 1B**). These counties, with the exception of Collier County, are mostly in the central part of the state, raising the question about the mechanisms underpinning this spatial expansion of local dengue risk beyond the southernmost counties of Florida, where most local dengue cases are reported (i.e., Miami-Dade & Monroe counties).

By modeling local dengue occurrence within Florida counties, we estimate that suitability for *Ae. aegypti*-borne transmission (index P) and the detection of travel-associated dengue cases are the most impactful on whether a county will report local cases in a given year (**Fig. 3**). Even when we controlled for those variables which could skew our data (e.g. 2022-2024 or Miami-Dade County), the impact of these drivers remained clear (**Fig. S3**). Throughout the timeseries, 2009 to 2024, we found that both transmission suitability and virus introductions via travelers were increasing — which are both likely tied to climate. Index P utilizes local temperature and humidity data and mosquito factors to estimate climate-driven transmission potential by *Ae. aegypti* mosquitoes that can be interpreted like the basic reproduction number (*R_0_* >1 transmission increasing, <1 decreasing) (*24*, *25*). We found that while index P is the highest in the southern part of the state, annual mean index P has been increasing throughout Florida (**Fig. 3, Fig. S3**). Meanwhile, increasing climate-driven dengue risk throughout the Americas is also increasing the rate of DENV introductions into Florida via travelers (*5*, *8*, *36–40*). While we did not conduct any predictive modeling, it is likely that the continuation of these climatic trends will not only lead to more frequent and larger outbreaks in the southern Miami-Dade and Monroe counties, but also further expand the detection of local cases northward.

## Limitations

DENV is known to cause asymptomatic or subclinical infections in 60-80% of infected people (*1*), which means that our data underestimates the true number of local cases. Local surveillance data is a function of case detection efforts, which can vary substantially between counties and over time. For example, no local dengue cases were reported in 2021. While this could be due to a true lack of cases, it is also possible that patients who presented with a self-limiting illness characterized by fever and other non-specific symptoms during the COVID-19 pandemic may not have sought healthcare or led providers to order the correct test. Likewise, our DENV sequencing and genomic surveillance efforts were not applied evenly during the study period and did not capture all of the major dengue events in Florida (e.g., 2009 and 2020 outbreaks in Monroe County, 2013 outbreak in Martin County). Sampling biases of DENV sequences from outside of Florida can lead to spurious identification of the source of introduction, especially if the true source is represented only by a few (or zero) DENV genomes in our global background dataset. Hence we limited our interpretations of introduction locations to regions (e.g. the Caribbean) unless other information supported more precise estimates (e.g. travel history showing DENV-3III_B.3.2 introductions from Cuba). Missing genomic data, from both within and outside of Florida, impacts some of our estimates of DENV lineage persistence (as discussed). Importantly, undersampling from outside Florida biases our results towards erroneously detecting trans-seasonal persistence, so we are confident that our conclusions are accurate in this case. While additional DENV sequencing from Florida will provide more clarity around the occurrence of trans-seasonal persistence, it would not change our conclusion for a lack of established endemic transmission cycle.

While *Ae. aegypti* mosquitoes are not the only DENV vector in Florida, we did not include *Ae. albopictus* transmission potential in our analysis. There has been significant debate about the comparative vectorial capacity of both DENV vectors (*1*, *41*, *42*). However, *Ae. aegypti* are strongly anthropophilic and are more likely to be found indoors; therefore many consider them to be better DENV vectors (*43*), since *Ae. albopictus* demonstrates opportunistic and zoophilic feeding behaviors. There is no continuous statewide, county-level mosquito surveillance system in Florida, but a state-wide survey from 2016 detected *Ae. albopictus* in all 56 counties included in the study (of the 67 total counties in the state), but only detected *Ae. aegypti* in 30 counties, all located in the central and southern part of the state (*44*). This was in agreement with subsequent species occurrence and abundance models that found a broad distribution for *Ae. albopictus*, with *Ae. aegypti* mostly restricted to southern Florida (*45*). Given that local dengue cases have not been reported in the northern panhandle, we decided to only focus on *Ae. aegypti* transmission potential and abundance. It is also possible that a prerequisite for the expansion of local dengue cases beyond central Florida is the northern expansion of *Ae. aegypti* mosquitoes.

## Conclusions

We used statistical modeling and genomic epidemiology to reveal the dynamics and drivers of local dengue cases in Florida. While we show that DENV transmission in Florida is dependent upon frequent virus introductions and it has not established local endemicity by 2024, we reveal that Florida is in a precarious situation with regards to future dengue risk. Local dengue cases are expanding to new counties and outbreaks are becoming more frequent, driven by climate factors that are both enhancing local transmission suitability and the frequency of introductions from increased activity in the region. Future research should focus on how to respond to the growing public health risk caused by dengue in Florida.

## Methods

### Ethics

The Institutional Review Boards (IRB) from the FDOH, Florida Gulf Coast University, and the Yale University Human Research Protection Program determined that pathogen genomic sequencing of de-identified remnant diagnostic samples as conducted in this study is not research involving human subjects (Yale IRB Protocol ID: 2000033281).

### Dengue case surveillance data

Dengue is a reportable disease in the United States. The FDOH detects cases through health care provider reporting, laboratory reporting, and syndromic surveillance. Weekly cumulative reports on local and travel-associated dengue case numbers were collected from 2009 to 2024 and are publicly available from the FDOH, including cases that were confirmed by both PCR and serologic assays (*7*). Travel-associated cases are defined as those that occurred in individuals who had traveled to a dengue-endemic country or territory within the two weeks prior to symptom onset. We aggregated the data by month and year of symptom onset and by location of diagnosis (i.e., Florida county).

### Sequencing and Genomic Dataset

We sequenced viral RNA from 133 local and 294 travel dengue cases detected in Florida, from serum collected between June 2010 through September 2024. Patient serum samples were processed using our DengueSeq protocol at Yale University or the FDOH as described previously (*8*, *46*). Briefly, RNA was isolated from each sample using the QIAamp Viral RNA Mini kit and were prepared for sequencing using the COVIDSeq RUO Kit (Illumina) using a custom primer scheme and modifications as previously described (*46*). cDNA was synthesized from RNA eluates and subsequently amplified in two separate highly multiplexed PCR reactions using primers designed for DENV 1 - 4. Amplified cDNA was combined, tagmented, purified, ligated to dual-index adapters, and pooled together for sequencing. At Yale, paired-end 150bp reads were generated on the Illumina NovaSeq at the Yale Center for Genomic Analysis, with 1 million reads ordered per library. Consensus genomes were generated at a coverage depth of 20X and filtered using a minimum frequency threshold of 0.75 in our DengueSeq bioinformatics pipeline using iVar (*46*, *47*), which is available at: https://github.com/grubaughlab/DENV-genomics. At FDOH, cDNA was first synthesized from RNA, after which amplicons were generated using the same custom primer scheme and modifications as described above (*46*). Libraries were prepared using the Nextera XT DNA Library Prep Kit (Illumina), and were subsequently sequenced on the in-house Illumina NextSeq 1000 and MiSeq instruments generating paired-end 251bp reads, targeting 250,000 reads (range of 150,000-400,000) per sample. GenBank accession numbers for all sequences that we generated in this study are available in **Table S2**.

To highlight dengue transmission dynamics into and within Florida, we filtered our global background dataset (n=15,766) to only include sequences from the same major lineages (n=5,153) as those detected in our locally acquired genomes. Major lineages offer greater genetic resolution than serotypes or genotypes and are useful in selecting epidemiologically-linked dengue genomic sequences (*48*). Lineage assignment was performed on all local, travel, and global dataset sequences using Nextclade (*48*, *49*). In total, our study analyzed 5,580 sequences collected between May 1961 through October 2024, spanning 89 countries. GenBank accession numbers for all 5,580 sequences can be found at: https://github.com/EmmaTS22/dengueFL.

### Phylogenetic analyses

All sequences were subject to a strict coverage cut-off of 70% genome coverage, and serotype-specific alignments were obtained from MAFFT v7.520, manually curated using Geneious v2022.1.1, and non-coding regions trimmed.. Using IQ-TREE v2.2.5, we generated an initial Maximum Likelihood (ML) tree with automatic model selection via ModelFinder (*50*). This initial ML tree was then inspected in TempEst v1.5.3 (*51*) to assess the molecular clock signal and to identify molecular clock outliers for removal. Sylvatic sequences were also excluded. Our final datasets consisted of DENV-1 (*n*=1688), DENV-2 (*n*=2438), DENV-3 (*n*=1093) and DENV-4 (*n*=361).

For computational efficiency and analytical flexibility, we split the analysis into topology estimation and phylogeographic inference. First, we estimated the topology in BEAST v1.10.5 (*52*) using a generalized time reversible (GTR) model (*53*), a gamma rate heterogeneity model with four rate categories (*54*), and a Hamiltonian Monte Carlo (HMC) random-effects relaxed clock model (*55*). The mean clock rate of each tree and their 95% highest posterior density CIs are: 9.90×10^−4^ (9.38×10^−4^ to 1.04×10^−3^), 1.24×10^−3^ (1.19×10^−3^ to 1.29×10^−3^), 1.08×10^−3^ (9.64×10^−4^ to 1.26×10^−3^), and 7.65×10^−4^ (6.40×10^−4^ to 9.09×10^−4^) substitutions/site/year for DENV-1 to -4 respectively.

To estimate effective population size dynamics, we used a Skygrid coalescent model (*56*) with a HMC operator (*57*). We defined the gridpoints externally to be at the start of each year, every year until 2000, then every 25 years until 1905, 1920, 1935, and 1940 for DENV-1 to -4 respectively. We chose these gridpoints to balance temporal resolution in more recent years with denser sampling against computational efficiency in older years. We placed final dates slightly before the estimated roots of the trees, as previously described (*58*). Depending on serotype, between two and three chains of 550m to 6,858m states were required for sufficiently high convergence and ESS values, with 33-72%% removed for burn-in.

To infer the geographic spread of each dengue virus serotype, we randomly sampled 500 trees from the post-burn-in posterior distribution as an empirical tree distribution as an input for phylogeographic discrete trait analysis (DTA) (*59*, *60*). Regions used as discrete traits in our DTA analysis were grouped with varying spatial resolutions to highlight transmission from the Americas into Florida. While sequences from Florida were grouped into counties (Miami-Dade, Polk, Pasco,..), other sequences from the Americas were grouped into their corresponding WHO regions (Caribbean, Central-America, North-America, South-America). All other sequences were grouped as ‘Global’.

In total, there were 16 unique regions used in our DTA analysis (DENV-1: 9 regions, DENV-2: 7 regions, DENV-3: 11 regions, DENV-4: 7 regions). Information on the number of sequences for each country, region, and DENV serotype can be found in Supplementary Table S1. We used an asymmetric discrete trait evolution model with a CTMC prior and logged complete Markov Jump histories. Each serotype analysis was run for two chains of 10m states, with 10% removed for burn-in. We assessed each analysis for convergence using Tracer v1.7.2 (*61*).

We inferred introductions of each serotype into Florida from the Maximum Clade Credibility (MCC) tree using a custom R script. We defined an introduction node as the oldest node in Florida where the direct ancestry was outside of Florida and counted all downstream nodes as part of the same introduction. The time of introduction was conservative, as was the time of the transition node itself, and should be interpreted as the latest approximate time that the introduction could have occurred. This is because including more samples can move the introduction node closer to the root of the tree (earlier in time), but not closer to the tip (later in time). The XML files used for all analyses can be found at: https://github.com/EmmaTS22/dengueFL.

We identified local transmission clusters as clades with three or more local DENV Florida sequences and we used the time to the most common recent ancestor (tMRCA) within Florida as a conservative date of introduction. Clades consisting of fewer samples resulted either from undersampling or the absence of epidemiological links (that would have otherwise generated larger clades). Therefore, those clades would not have been sufficiently informative of trans-seasonal persistence, since their long branch lengths would lead to introduction time estimates with poor temporal resolution (wide 95% highest posterior density CIs).

We characterized transmission routes into and within Florida using the posterior distribution of the complete Markov Jump history. Each Markov Jump describes the direction of movement between two traits and its corresponding time. Using the BSSVS (Bayesian stochastic search variable selection) approach implemented in BEAST v1.10.5, we visualized only viral lineage movements into or out of Florida that were statistically supported (Bayes factor >3, posterior probability > 0.5) (*60*, *62*).

### Mosquito transmission suitability index

We estimated the local potential for DENV transmission by *Ae. aegypti* using a previously published suitability measure: index P (*24*, *25*). Index P estimates the suitability for transmission of mosquito-borne viruses using meteorological variables, such as temperature and relative humidity, as well as epi-entomological parameters, such as biting rate and extrinsic incubation period. We combined daily meteorological data with previously-documented ecological and entomological priors (**Table S6**) using the R package MVSE (*25*) to estimate daily transmission potential. Index P estimates were aggregated to calculate the mean and maximum values, as well as total days with index P > 0 (transmission was possible) at both annual and monthly resolutions. These annual estimates were used to model the annual occurrence of local dengue cases in Florida counties, and monthly estimates were used to model monthly dengue incidence in Miami-Dade County.

### Socioeconomic and climate variables

As a proxy for socioeconomic vulnerability, we calculated the SVI for each county and year as described by the CDC, where higher values correspond to poverty, lack of accessible transportation, and more crowded housing (*63*). We used human population density as a proxy for urbanization, which we calculated on a county level using data from the U.S. Census Bureau (https://www.census.gov). To examine the potential impact of meteorological variables on the occurrence of local dengue cases in Floridian counties, we included temperature (°C), precipitation (mm), and relative humidity (%) in the model. For each county, we obtained daily weather data from Visual Crossing Weather (https://www.visualcrossing.com/). From these daily estimates, we calculated the yearly mean, minimum and maximum temperature, total and mean precipitation, days without precipitation, and mean relative humidity to model the annual occurrence of local dengue cases at the county level in Florida. In a similar fashion, these daily estimates were also aggregated to monthly values to model the monthly incidence of local dengue cases in Miami-Dade County.

### Vector abundance

Miami-Dade County’s standardized mosquito surveillance system was established in response to its 2016 Zika outbreak, as previously described (*29*). *Ae. aegypti* is the primary mosquito vector in the county. To model the impact of vector abundance on monthly DENV incidence in Miami-Dade, we obtained *Ae. aegypti* trap data from the Miami-Dade County Mosquito Control Division for 2017-2024, during which most DENV cases in Miami-Dade were detected (94% of the total number of cases, n=310). During this time period, 99804 mosquito traps (86,181 BG-Sentinel 2 traps and 13,623 CDC light traps) were set weekly and baited with CO2 (*64*). All mosquitoes were morphologically identified by the Miami-Dade County Mosquito Control Laboratory. Male mosquitoes are not attracted to CO2-based traps and were considered accidental catches and excluded from our analyses. We aggregated the data (cumulative *Ae. aegypti*-per-trap) by month and included this in our monthly incidence model for Miami-Dade County.

### Statistical Methods

To infer the effects of our covariates (e.g., travel cases, mosquito abundance) on local dengue risk in Florida, we fitted spatiotemporal models in a Bayesian framework using the R package ‘R-INLA’ to implement integrated nested Laplace approximation (INLA v23.09.09). The INLA framework is a computationally tractable alternative to MCMC as it approximates the marginal posterior distributions for all parameters without the need for posterior sampling. We have used the default penalized complexity priors from INLA for the spatial random effects occurrence and incidence model, and we have set uninformative Gaussian priors for fixed effects.

### Occurrence Model

For **Fig. 3**, we modeled the annual occurrence of local dengue cases in Florida for county *i* and year *t* (*Y_i,t_*) using logistic regression such that

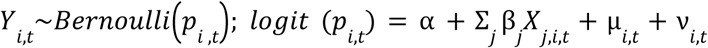

where α is the intercept, 𝑋_𝑗,𝑖,𝑡_ is a matrix of values for the *j*–th covariate with linear fixed effects β*_j_*, µ_𝑖,𝑡_ and ν_𝑖,𝑡_ are the spatially structured and unstructured random effects at time period t, jointly specified as a BYM model (*28*, *65*).

The BYM model accounts for spatial correlation in local dengue cases between counties, using random effects (µ_𝑖,𝑡_ and ν_𝑖,𝑡_) to capture unexplained spatial variation in the annual occurrence of local dengue cases and how observations between neighboring counties are potentially more similar than those between more distant counties. Our baseline model included only these random effects (µ_𝑖,𝑡_ outcome in that year. and ν_𝑖,𝑡_), which produced a smooth version of occurrence for the overall outcome in that year.

Next, we added covariates individually to our baseline model to examine their effects on annual occurrence. We only kept covariates that lowered the WAIC in our visualization of their posterior marginal distributions. This left us with the following covariates: climate (annual minimum, mean, and maximum temperature, number of days in a year without precipitation, annual mean and cumulative precipitation, annual mean relative humidity), index P (annual mean and maximum index P, number of days in a year with index P > 0), population density, SVI, and travel cases (annual travel case occurrence, total annual travel cases, and first month travel cases were detected).

### Incidence Model

For **Fig. 4**, we modeled the monthly case counts of local dengue in Miami-Dade County (*Y_t_*) with a Poisson regression such that

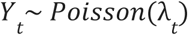

where we modeled the expected number of cases for Miami-Dade County during month *t* (λ*_t_*) with the following linear predictor

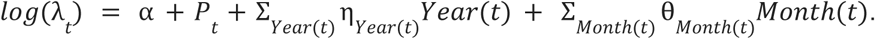

Here, α is the intercept and 𝑃_𝑡_ is log population included as an offset term. To account for unexplained variation in local dengue risk between years and months, we included fixed effects η_𝑌𝑒𝑎𝑟(𝑡)_ (for years 2009 to 2024) and θ_𝑀𝑜𝑛𝑡ℎ(𝑡)_ (for months January to December) acting on indicator variables *Year*(*t*) and *Month*(*t*) which take on values 0 or 1 depending on the corresponding year and month for month *t*.

Fitting this fixed effects-only model as a baseline, we conduct model selection to develop a multi-variable model incorporating covariates from the following broad categories: temperature, hydrometeorological, travel cases, mosquito abundance, and index P (**Table S5**). First, we chose each covariate’s best-fitting type (e.g., either monthly mean, minimum, or maximum values for temperature) and functional form (e.g., either linear or non-linear) by adding each covariate to the baseline model and compared their WAIC (*66*). Then, from each covariate category, we chose the covariate type and functional form with the lowest WAIC for inclusion into our full multivariable model (**Table S7**). Between all selected covariates, we assessed their multicollinearity by calculating their variance inflation factors (VIF), and considered VIF ≥ 5 as evidence of high collinearity. Covariates selected for our full-fit model are: travel cases, number of *Ae. aegypti* mosquitoes per trap, maximum temperature, mean humidity, and mean index P. The best-fitting form for all aforementioned covariates was non-linear, which was modeled as a second-order random walk.

In addition, we tested another model using a negative binomial likelihood to account for overdispersion, incorporating the same yearly and monthly fixed effects. Ultimately, the negative binomial full-fit model performed worse (WAIC 338) than the Poisson full-fit model (WAIC 331).

## Data availability

GenBank accession numbers for all sequences that we generated in this study are available in **Table S2**. Covariates we used for modelling the annual occurrence and monthly incidence of local DENV cases are summarized in **Table S5**.

## Code availability

All code and model results are available at: https://github.com/EmmaTS22/dengueFL.

## Acknowledgements

We acknowledge Olga Ospina and Hunter Scott from the Bureau of Epidemiology at FDOH and D. Weinberger, S. Taylor, and P. Jack for technical discussions. This publication was made possible by the National Institute of Allergy and Infectious Diseases of the National Institutes of Health (NIH) under Award Number DP2AI176740 (NDG), pilot funds from the Yale Planetary Sciences via the Three Cairns Climate Impact Innovation Fund and the Office of the Provost AI Initiatives (NDG), the National Institute of General Medical Sciences R21GM142011 (SFM), the National Science Scholarship from the Agency for Science, Technology, and Research (A*STAR) (YTC), Singapore. The findings and conclusions in this report are those of the author(s) and do not necessarily represent the official position of the NIH, FDOH, or A*STAR.

## Author Contributions

E.T.S, Y.T.C, R.L. and N.D.G. designed the study and wrote the paper. A.D., P.C., S.S., V.M., B.S., R.Z., D.S., and A.M. collected the surveillance data. C.V. and M.Mo. collected the mosquito data. T.L., E.K., J.V., M.Mi., L.M.P., S.F.M., M.I.B., C.B.F.V., and L.H. sequenced the samples and performed bioinformatic analyses. E.T.S., Y.T.C., R.L., J.L.W., C.J.C, and V.H. performed statistical and phylogenetic analyses. V.H., A.M., and N.D.G. supervised the project. All authors reviewed and approved the final version of the manuscript.

## Supplemental Figures & Tables

**Supplemental Figure 1.**
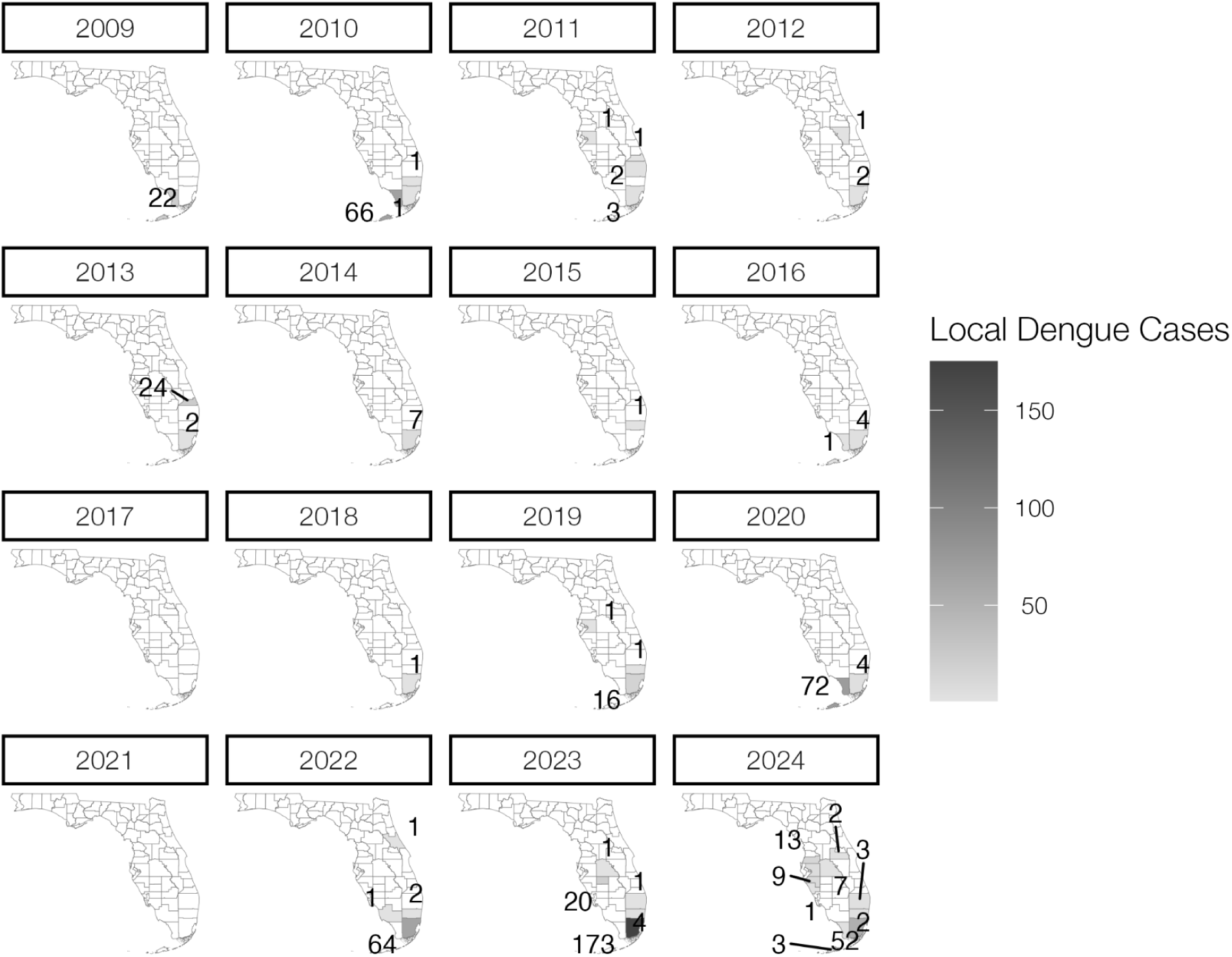
Total number of local dengue cases per county reported in Florida from 2009 to 2024. There were no local cases reported in 2017 or 2021.

**Supplemental Figure 2.**
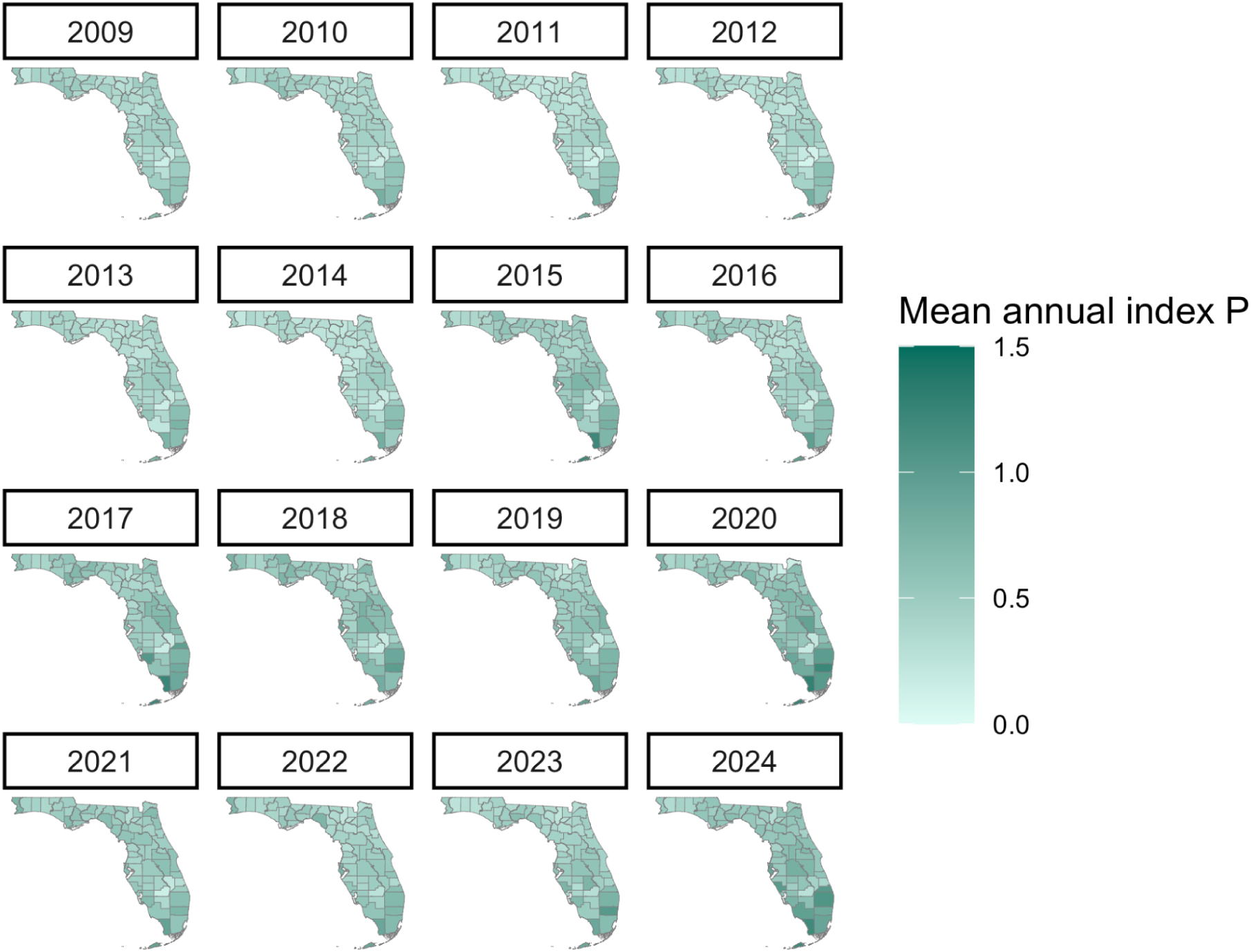
Yearly estimated mean index P for DENV transmitted by *Aedes aegypti* mosquitoes per county in Florida from 2009 to 2024.

**Supplemental Figure 3.**
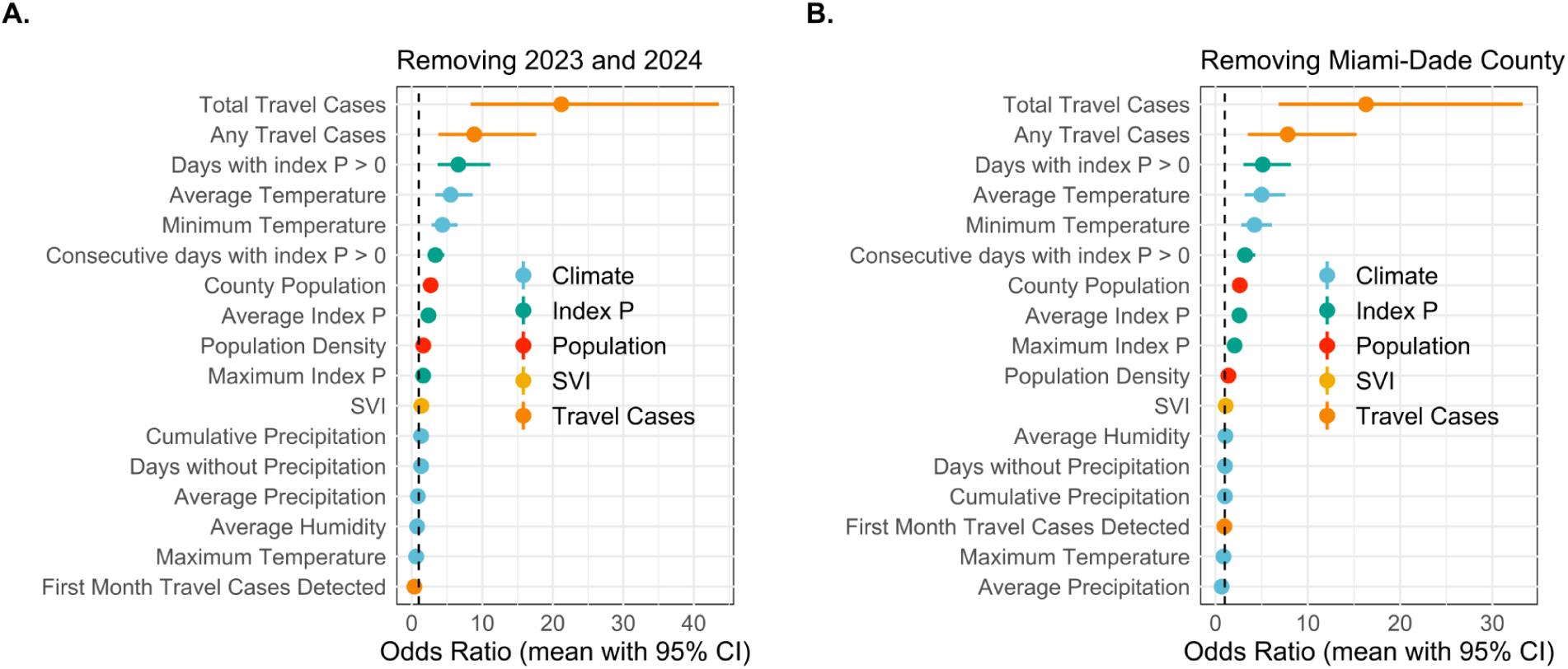
Sensitivity analysis to understand the marginal linear fixed effects of annual local dengue case occurrence by county by various covariates if (A) the 2023-2024 are removed from the analysis or (B) Miami-Dade County were removed from the analysis. The circles indicate the predicted mean and the lines indicate the 95% confidence interval. Covariates are colored by category.

**Table S1.** Genomic dataset including all travel-associated (n=294) and locally acquired (n=133) dengue cases reported by the FDOH from 2009-2024, and a global background dataset (n=5,153) filtered for major lineages matching those of locally acquired dengue cases. Sequence counts are broken down by sample collection location and serotype. Sample collection locations are grouped into traits for phylogeographic inference (Region(DTA)).

| Location | Region (DTA) | DENV-1 | DENV-2 | DENV-3 | DENV-4 |
| --- | --- | --- | --- | --- | --- |
| Florida (USA) | Alachua | 0 | 1 | 0 | 0 |
| Florida (USA) | Hardee | 1 | 0 | 0 | 0 |
| Florida (USA) | Hillsborough | 0 | 0 | 1 | 0 |
| Florida (USA) | Manatee | 0 | 0 | 0 | 1 |
| Florida (USA) | Martin | 1 | 0 | 0 | 0 |
| Florida (USA) | Miami-Dade | 1 | 14 | 95 | 2 |
| Florida (USA) | Monroe | 12 | 0 | 0 | 0 |
| Florida (USA) | Orange | 0 | 0 | 1 | 0 |
| Florida (USA) | Palm-Beach | 0 | 0 | 1 | 0 |
| Florida (USA) | Pasco | 0 | 0 | 1 | 0 |
| Florida (USA) | Polk | 0 | 0 | 1 | 0 |
| Barbados | Caribbean | 0 | 1 | 0 | 0 |
| Cuba | Caribbean | 23 | 98 | 293 | 48 |
| Dominica | Caribbean | 0 | 0 | 0 | 2 |
| Dominican-Republic | Caribbean | 8 | 49 | 68 | 2 |
| Guadeloupe | Caribbean | 4 | 50 | 2 | 0 |
| Haiti | Caribbean | 6 | 3 | 2 | 5 |
| Jamaica | Caribbean | 2 | 9 | 10 | 1 |
| Martinique | Caribbean | 0 | 55 | 7 | 0 |
| Puerto-Rico | Caribbean | 26 | 11 | 13 | 29 |
| Saint-Barthelemy | Caribbean | 1 | 4 | 0 | 0 |
| Saint-Croix | Caribbean | 0 | 1 | 0 | 0 |
| Saint-Kitts-and-Nevis | Caribbean | 0 | 1 | 0 | 0 |
| Saint-Lucia | Caribbean | 0 | 0 | 2 | 0 |
| Saint-Martin | Caribbean | 3 | 1 | 0 | 0 |
| Trinidad-and-Tobago | Caribbean | 0 | 1 | 7 | 0 |
| US-Virgin-Islands | Caribbean | 0 | 1 | 0 | 0 |
| Belize | Central-America | 0 | 1 | 0 | 0 |
| Costa-Rica | Central-America | 2 | 1 | 5 | 0 |
| El-Salvador | Central-America | 0 | 0 | 2 | 2 |
| Guatemala | Central-America | 0 | 3 | 8 | 0 |
| Honduras | Central-America | 1 | 5 | 13 | 0 |
| Nicaragua | Central-America | 63 | 289 | 0 | 6 |
| Panama | Central-America | 0 | 1 | 1 | 2 |
| Angola | Global | 0 | 0 | 1 | 0 |
| Australia | Global | 0 | 2 | 2 | 0 |
| Bangladesh | Global | 2 | 8 | 36 | 0 |
| Benin | Global | 1 | 0 | 0 | 0 |
| Bhutan | Global | 0 | 0 | 44 | 0 |
| Brunei | Global | 1 | 2 | 0 | 0 |
| Burkina-Faso | Global | 6 | 0 | 8 | 0 |
| Cambodia | Global | 1 | 114 | 1 | 0 |
| Cameroon | Global | 0 | 1 | 0 | 0 |
| China | Global | 109 | 277 | 75 | 0 |
| Djibouti | Global | 1 | 0 | 0 | 0 |
| Egypt | Global | 0 | 1 | 0 | 0 |
| Eritrea | Global | 1 | 0 | 0 | 0 |
| Ethiopia | Global | 0 | 0 | 2 | 0 |
| France | Global | 1 | 4 | 0 | 0 |
| Gabon | Global | 0 | 0 | 5 | 0 |
| Ghana | Global | 1 | 0 | 0 | 0 |
| Guam | Global | 0 | 1 | 0 | 0 |
| India | Global | 73 | 26 | 59 | 1 |
| Indonesia | Global | 0 | 10 | 18 | 4 |
| Israel | Global | 2 | 0 | 2 | 0 |
| Italy | Global | 7 | 0 | 3 | 0 |
| Ivory-Coast | Global | 3 | 0 | 0 | 0 |
| Kenya | Global | 5 | 0 | 5 | 0 |
| Kiribati | Global | 0 | 0 | 0 | 1 |
| Malaysia | Global | 2 | 43 | 4 | 0 |
| Maldives | Global | 0 | 1 | 4 | 0 |
| Mauritania | Global | 1 | 0 | 0 | 0 |
| Mayotte | Global | 31 | 0 | 0 | 0 |
| Myanmar | Global | 0 | 0 | 39 | 0 |
| Nepal | Global | 4 | 23 | 2 | 0 |
| Niger | Global | 0 | 0 | 1 | 0 |
| Nigeria | Global | 3 | 0 | 0 | 0 |
| Niue | Global | 0 | 0 | 0 | 1 |
| Pakistan | Global | 23 | 14 | 6 | 0 |
| Papua-New-Guinea | Global | 0 | 0 | 5 | 0 |
| Philippines | Global | 0 | 2 | 0 | 0 |
| Reunion | Global | 43 | 5 | 17 | 0 |
| Saudi-Arabia | Global | 0 | 0 | 1 | 0 |
| Senegal | Global | 3 | 0 | 12 | 1 |
| Seychelles | Global | 1 | 0 | 0 | 0 |
| Singapore | Global | 102 | 124 | 14 | 2 |
| South-Korea | Global | 0 | 1 | 0 | 0 |
| Sri-Lanka | Global | 39 | 156 | 58 | 0 |
| Taiwan | Global | 0 | 1 | 0 | 0 |
| Tanzania | Global | 2 | 0 | 0 | 0 |
| Thailand | Global | 0 | 13 | 60 | 0 |
| Timor-Leste | Global | 0 | 0 | 4 | 0 |
| UAE | Global | 0 | 1 | 0 | 0 |
| Vietnam | Global | 0 | 50 | 0 | 0 |
| Arizona (USA) | North-America | 0 | 0 | 3 | 0 |
| California (USA) | North-America | 1 | 5 | 28 | 0 |
| Mexico | North-America | 127 | 34 | 12 | 4 |
| Texas (USA) | North-America | 1 | 5 | 0 | 0 |
| Argentina | South-America | 20 | 1 | 0 | 0 |
| Bolivia | South-America | 3 | 1 | 0 | 1 |
| Brazil | South-America | 753 | 602 | 9 | 87 |
| Colombia | South-America | 19 | 121 | 1 | 12 |
| Ecuador | South-America | 16 | 33 | 0 | 2 |
| French-Guiana | South-America | 0 | 12 | 6 | 0 |
| Guyana | South-America | 0 | 0 | 2 | 0 |
| Paraguay | South-America | 40 | 37 | 0 | 98 |
| Peru | South-America | 51 | 59 | 10 | 0 |
| Suriname | South-America | 0 | 2 | 0 | 0 |
| Venezuela | South-America | 35 | 46 | 1 | 47 |

**Table S2.** Genbank accession numbers, collection dates & regions, and lineage information for sequences sampled from either local (Local=1, n=133) or travel-associated (Travel=1, n=294) dengue cases in Florida.

| GenBank Accession | Year | Month | Day | Local | Travel | Location | Region (DTA) | Serotype | Lineage |
| --- | --- | --- | --- | --- | --- | --- | --- | --- | --- |
| OQ821370 | 2010 | 6 | 24 | 1 | 0 | Florida (USA) | Monroe | 1 | 1V_C |
| OQ821371 | 2010 | 6 | 30 | 1 | 0 | Florida (USA) | Monroe | 1 | 1V_C |
| OQ821353 | 2010 | 7 | 28 | 0 | 1 | Dominican-Republic | Caribbean (Travel) | 1 | 1V_E |
| OQ821494 | 2010 | 7 | 30 | 0 | 1 | Puerto-Rico | Caribbean (Travel) | 2 | 2III_B |
| OQ821632 | 2010 | 8 | 5 | 0 | 1 | Puerto-Rico | Caribbean (Travel) | 4 | 4II_B |
| OQ821372 | 2010 | 8 | 6 | 1 | 0 | Florida (USA) | Monroe | 1 | 1V_C |
| OQ821373 | 2010 | 8 | 11 | 1 | 0 | Florida (USA) | Monroe | 1 | 1V_C |
| OQ821374 | 2010 | 8 | 27 | 1 | 0 | Florida (USA) | Monroe | 1 | 1V_C |
| OQ821375 | 2010 | 10 | 11 | 1 | 0 | Florida (USA) | Monroe | 1 | 1V_C |
| JQ675358 | 2010 | 10 |  | 1 | 0 | Florida (USA) | Monroe | 1 | 1V_C |
| OQ821376 | 2010 | 11 | 20 | 1 | 0 | Florida (USA) | Monroe | 1 | 1V_C |
| OQ821628 | 2012 | 8 | 20 | 0 | 1 | Cuba | Caribbean (Travel) | 4 | 4II_B.1 |
| OQ821364 | 2012 | 9 | 2 | 0 | 1 | Jamaica | Caribbean (Travel) | 1 | 1V_E |
| OQ821629 | 2012 | 10 | 3 | 0 | 1 | Cuba | Caribbean (Travel) | 4 | 4II_B.1 |
| OM833059 | 2013 | 7 | 24 | 1 | 0 | Florida (USA) | Monroe | 1 | 1V_E.3 |
| OQ821650 | 2013 | 8 | 21 | 0 | 1 | Dominica | Caribbean (Travel) | 4 | 4II_A.1 |
| OQ821377 | 2013 | 8 | 29 | 1 | 0 | Florida (USA) | Martin | 1 | 1V_E.2 |
| OQ821633 | 2013 | 9 | 6 | 0 | 1 | Haiti | Caribbean (Travel) | 4 | 4II_B |
| OQ821354 | 2013 | 9 | 21 | 0 | 1 | Dominican-Republic | Caribbean (Travel) | 1 | 1V_E.2 |
| OQ821383 | 2013 | 12 | 1 | 0 | 1 | Cuba | Caribbean (Travel) | 2 | 2III_D.1.2 |
| OQ821626 | 2013 | 12 | 6 | 0 | 1 | Cuba | Caribbean (Travel) | 4 | 4II_B.1 |
| OQ821472 | 2014 | 1 | 12 | 0 | 1 | Dominican-Republic | Caribbean (Travel) | 2 | 2III_C.1 |
| OQ821630 | 2014 | 3 | 13 | 0 | 1 | Bolivia | South-America (Travel) | 4 | 4II_B.1.1 |
| OQ821612 | 2014 | 7 | 2 | 1 | 0 | Florida (USA) | Miami-Dade | 3 | 3I_A |
| OQ821627 | 2014 | 8 | 6 | 0 | 1 | Cuba | Caribbean (Travel) | 4 | 4II_B.1 |
| OQ821476 | 2014 | 10 | 14 | 0 | 1 | Honduras | Central-America (Travel) | 2 | 2III_D.1.2 |
| OQ821327 | 2015 | 5 | 19 | 0 | 1 | Brazil | South-America (Travel) | 1 | 1V_F |
| OQ821500 | 2015 | 8 | 31 | 0 | 1 | Venezuela | South-America (Travel) | 2 | 2III_D.2 |
| OQ821475 | 2015 | 10 | 28 | 0 | 1 | Haiti | Caribbean (Travel) | 2 | 2III_C.1.2 |
| OQ821503 | 2016 | 1 | 13 | 0 | 1 | Cuba | Caribbean (Travel) | 3 | 3III_C.1 |
| OQ821501 | 2016 | 1 | 23 | 0 | 1 | Venezuela | South-America (Travel) | 2 | 2III_D.2 |
| OQ821625 | 2016 | 2 | 5 | 0 | 1 | Cuba | Caribbean (Travel) | 4 | 4II_B.1 |
| OQ821328 | 2016 | 2 | 8 | 0 | 1 | Brazil | South-America (Travel) | 1 | 1V_A |
| OQ821332 | 2016 | 3 | 15 | 0 | 1 | Costa-Rica | Central-America (Travel) | 1 | 1V_C |
| OQ821649 | 2016 | 3 | 21 | 0 | 1 | Jamaica | Caribbean (Travel) | 4 | 4II_A.1 |
| KX702404 | 2016 | 5 | 22 | 1 | 0 | Florida (USA) | Alachua | 2 | 2III_C |
| OQ821601 | 2016 | 6 | 11 | 0 | 1 | Jamaica | Caribbean (Travel) | 3 | 3III_B.3.1 |
| OQ821331 | 2016 | 7 | 12 | 0 | 1 | Colombia | South-America (Travel) | 1 | 1V_D.1 |
| OQ821602 | 2016 | 7 | 27 | 0 | 1 | Jamaica | Caribbean (Travel) | 3 | 3III_B.3.1 |
| OQ821381 | 2016 | 7 | 28 | 0 | 1 | Colombia | South-America (Travel) | 2 | 2III_D.3 |
| OQ821382 | 2016 | 8 | 1 | 0 | 1 | Costa-Rica | Central-America (Travel) | 2 | 2III_D.1.1 |
| OQ821603 | 2016 | 9 | 12 | 0 | 1 | Jamaica | Caribbean (Travel) | 3 | 3III_B.3.1 |
| MN192436 | 2016 |  |  | 1 | 0 | Florida (USA) | Manatee | 4 | 4II_B |
| OQ821474 | 2017 | 4 | 22 | 0 | 1 | Guatemala | Central-America (Travel) | 2 | 2III_D.1.2 |
| OQ821384 | 2017 | 6 | 29 | 0 | 1 | Cuba | Caribbean (Travel) | 2 | 2III_C.2 |
| OQ821480 | 2017 | 9 | 8 | 0 | 1 | India | South-Asia (Travel) | 2 | 2II_A.2.1 |
| OQ821360 | 2018 | 7 | 5 | 0 | 1 | Haiti | Caribbean (Travel) | 1 | 1V_E.2 |
| OQ821385 | 2018 | 11 | 4 | 0 | 1 | Cuba | Caribbean (Travel) | 2 | 2III_C.2 |
| OQ821386 | 2018 | 11 | 20 | 0 | 1 | Cuba | Caribbean (Travel) | 2 | 2III_C.2 |
| OQ821387 | 2018 | 12 | 5 | 0 | 1 | Cuba | Caribbean (Travel) | 2 | 2III_C.2 |
| OQ821388 | 2019 | 1 | 6 | 0 | 1 | Cuba | Caribbean (Travel) | 2 | 2III_C.2 |
| OQ821379 | 2019 | 1 | 7 | 0 | 1 | Venezuela | South-America (Travel) | 1 | 1V_D.1 |
| OQ821389 | 2019 | 1 | 13 | 0 | 1 | Cuba | Caribbean (Travel) | 2 | 2III_C.2 |
| OQ821604 | 2019 | 1 | 15 | 0 | 1 | Jamaica | Caribbean (Travel) | 3 | 3III_B.3.1 |
| OQ821605 | 2019 | 2 | 1 | 0 | 1 | Jamaica | Caribbean (Travel) | 3 | 3III_B.3.1 |
| OQ821390 | 2019 | 3 | 20 | 0 | 1 | Cuba | Caribbean (Travel) | 2 | 2III_C.2 |
| OQ821391 | 2019 | 3 | 20 | 0 | 1 | Cuba | Caribbean (Travel) | 2 | 2III_C.2 |
| OQ821392 | 2019 | 6 | 3 | 0 | 1 | Cuba | Caribbean (Travel) | 2 | 2III_D.1.1 |
| OQ821486 | 2019 | 6 | 6 | 0 | 1 | Nicaragua | Central-America (Travel) | 2 | 2III_D.1.2 |
| OQ821393 | 2019 | 6 | 7 | 0 | 1 | Cuba | Caribbean (Travel) | 2 | 2III_C.2 |
| OQ821394 | 2019 | 6 | 29 | 0 | 1 | Cuba | Caribbean (Travel) | 2 | 2III_C.2 |
| OQ821355 | 2019 | 7 | 4 | 0 | 1 | Dominican-Republic | Caribbean (Travel) | 1 | 1V_E.2 |
| OQ821395 | 2019 | 7 | 10 | 0 | 1 | Cuba | Caribbean (Travel) | 2 | 2III_C.2 |
| OQ821396 | 2019 | 7 | 10 | 0 | 1 | Cuba | Caribbean (Travel) | 2 | 2III_C.2 |
| OQ821397 | 2019 | 7 | 11 | 0 | 1 | Cuba | Caribbean (Travel) | 2 | 2III_C.2 |
| OQ821496 | 2019 | 7 | 11 | 1 | 0 | Florida (USA) | Miami-Dade | 2 | 2III_D.1.1 |
| OQ821477 | 2019 | 7 | 12 | 0 | 1 | Honduras | Central-America (Travel) | 2 | 2III_D.1.2 |
| OQ821487 | 2019 | 7 | 14 | 0 | 1 | Nicaragua | Central-America (Travel) | 2 | 2III_D.1.2 |
| OQ821398 | 2019 | 7 | 15 | 0 | 1 | Cuba | Caribbean (Travel) | 2 | 2III_C.2 |
| OQ821606 | 2019 | 7 | 15 | 0 | 1 | Jamaica | Caribbean (Travel) | 3 | 3III_B.3.1 |
| OQ821330 | 2019 | 7 | 17 | 0 | 1 | Cambodia | Southeast-Asia (Travel) | 1 | 1I_E.1 |
| OQ821399 | 2019 | 7 | 21 | 0 | 1 | Cuba | Caribbean (Travel) | 2 | 2III_C.2 |
| OQ821400 | 2019 | 7 | 22 | 0 | 1 | Cuba | Caribbean (Travel) | 2 | 2III_C.2 |
| OQ821401 | 2019 | 7 | 24 | 0 | 1 | Cuba | Caribbean (Travel) | 2 | 2III_C.2 |
| OQ821333 | 2019 | 7 | 26 | 0 | 1 | Cuba | Caribbean (Travel) | 1 | 1V_D.1.1 |
| OQ821402 | 2019 | 7 | 27 | 0 | 1 | Cuba | Caribbean (Travel) | 2 | 2III_C.2 |
| OQ821403 | 2019 | 7 | 27 | 0 | 1 | Cuba | Caribbean (Travel) | 2 | 2III_C.2 |
| OQ821404 | 2019 | 7 | 28 | 0 | 1 | Cuba | Caribbean (Travel) | 2 | 2III_C.2 |
| OQ821493 | 2019 | 7 | 31 | 0 | 1 | Philippines | Southeast-Asia (Travel) | 2 | 2II_C.1 |
| OQ821405 | 2019 | 8 | 1 | 0 | 1 | Cuba | Caribbean (Travel) | 2 | 2III_C.2 |
| OQ821478 | 2019 | 8 | 2 | 0 | 1 | Honduras | Central-America (Travel) | 2 | 2III_D.1.2 |
| OQ821406 | 2019 | 8 | 3 | 0 | 1 | Cuba | Caribbean (Travel) | 2 | 2III_C.2 |
| OQ821407 | 2019 | 8 | 3 | 0 | 1 | Cuba | Caribbean (Travel) | 2 | 2III_D.1.1 |
| OQ821408 | 2019 | 8 | 6 | 0 | 1 | Cuba | Caribbean (Travel) | 2 | 2III_C.2 |
| OQ821356 | 2019 | 8 | 12 | 0 | 1 | Dominican-Republic | Caribbean (Travel) | 1 | 1V_E.2 |
| OQ821361 | 2019 | 8 | 12 | 0 | 1 | Haiti | Caribbean (Travel) | 1 | 1V_E.2 |
| OQ821607 | 2019 | 8 | 12 | 0 | 1 | Jamaica | Caribbean (Travel) | 3 | 3III_B.3.1 |
| OQ821488 | 2019 | 8 | 12 | 0 | 1 | Nicaragua | Central-America (Travel) | 2 | 2III_D.1.2 |
| OQ821362 | 2019 | 8 | 14 | 0 | 1 | Haiti | Caribbean (Travel) | 1 | 1V_E.2 |
| OQ821334 | 2019 | 8 | 18 | 0 | 1 | Cuba | Caribbean (Travel) | 1 | 1V_D.1.1 |
| OQ821483 | 2019 | 8 | 19 | 0 | 1 | Mexico | North-America (Travel) | 2 | 2III_D.1.2 |
| OQ821409 | 2019 | 8 | 22 | 0 | 1 | Cuba | Caribbean (Travel) | 2 | 2III_C.2 |
| OQ821363 | 2019 | 8 | 23 | 0 | 1 | Honduras | Central-America (Travel) | 1 | 1V_C |
| OQ821410 | 2019 | 8 | 25 | 0 | 1 | Cuba | Caribbean (Travel) | 2 | 2III_C.2 |
| OQ821411 | 2019 | 8 | 26 | 0 | 1 | Cuba | Caribbean (Travel) | 2 | 2III_C.2 |
| OQ821412 | 2019 | 8 | 26 | 0 | 1 | Cuba | Caribbean (Travel) | 2 | 2III_C.2 |
| OQ821479 | 2019 | 8 | 27 | 0 | 1 | Honduras | Central-America (Travel) | 2 | 2III_D.1.2 |
| OQ821489 | 2019 | 8 | 27 | 0 | 1 | Nicaragua | Central-America (Travel) | 2 | 2III_D.1.2 |
| OQ821335 | 2019 | 8 | 28 | 0 | 1 | Cuba | Caribbean (Travel) | 1 | 1V_E.2 |
| OQ821413 | 2019 | 8 | 28 | 0 | 1 | Cuba | Caribbean (Travel) | 2 | 2III_C.2 |
| OQ821497 | 2019 | 8 | 29 | 1 | 0 | Florida (USA) | Miami-Dade | 2 | 2III_D.1.2 |
| OQ821414 | 2019 | 8 | 30 | 0 | 1 | Cuba | Caribbean (Travel) | 2 | 2III_C.2 |
| OQ821415 | 2019 | 8 | 30 | 0 | 1 | Cuba | Caribbean (Travel) | 2 | 2III_C.2 |
| OQ821416 | 2019 | 9 | 2 | 0 | 1 | Cuba | Caribbean (Travel) | 2 | 2III_C.2 |
| OQ821417 | 2019 | 9 | 4 | 0 | 1 | Cuba | Caribbean (Travel) | 2 | 2III_C.2 |
| OQ821490 | 2019 | 9 | 5 | 0 | 1 | Nicaragua | Central-America (Travel) | 2 | 2III_D.1.2 |
| OQ821418 | 2019 | 9 | 7 | 0 | 1 | Cuba | Caribbean (Travel) | 2 | 2III_C.2 |
| OQ821419 | 2019 | 9 | 8 | 0 | 1 | Cuba | Caribbean (Travel) | 2 | 2III_D.1.1 |
| OQ821420 | 2019 | 9 | 8 | 0 | 1 | Cuba | Caribbean (Travel) | 2 | 2III_D.1.1 |
| OQ821357 | 2019 | 9 | 10 | 0 | 1 | Dominican-Republic | Caribbean (Travel) | 1 | 1V_E.2 |
| OQ821421 | 2019 | 9 | 13 | 0 | 1 | Cuba | Caribbean (Travel) | 2 | 2III_C.2 |
| OQ821422 | 2019 | 9 | 13 | 0 | 1 | Cuba | Caribbean (Travel) | 2 | 2III_C.2 |
| OQ821498 | 2019 | 9 | 14 | 1 | 0 | Florida (USA) | Miami-Dade | 2 | 2III_D.1.2 |
| OQ821424 | 2019 | 9 | 16 | 0 | 1 | Cuba | Caribbean (Travel) | 2 | 2III_C.2 |
| OQ821425 | 2019 | 9 | 18 | 0 | 1 | Cuba | Caribbean (Travel) | 2 | 2III_C.2 |
| OQ821426 | 2019 | 9 | 18 | 0 | 1 | Cuba | Caribbean (Travel) | 2 | 2III_C.2 |
| OQ821428 | 2019 | 9 | 18 | 0 | 1 | Cuba | Caribbean (Travel) | 2 | 2III_D.1.1 |
| OQ821429 | 2019 | 9 | 20 | 0 | 1 | Cuba | Caribbean (Travel) | 2 | 2III_D.1.1 |
| OQ821378 | 2019 | 9 | 20 | 1 | 0 | Florida (USA) | Miami-Dade | 1 | 1III_A.3 |
| OQ821430 | 2019 | 9 | 21 | 0 | 1 | Cuba | Caribbean (Travel) | 2 | 2III_C.2 |
| OQ821431 | 2019 | 9 | 23 | 0 | 1 | Cuba | Caribbean (Travel) | 2 | 2III_C.2 |
| OQ821432 | 2019 | 9 | 23 | 0 | 1 | Cuba | Caribbean (Travel) | 2 | 2III_C.2 |
| OQ821433 | 2019 | 9 | 24 | 0 | 1 | Cuba | Caribbean (Travel) | 2 | 2III_C.2 |
| OQ821434 | 2019 | 9 | 26 | 0 | 1 | Cuba | Caribbean (Travel) | 2 | 2III_D.1.1 |
| OQ821435 | 2019 | 9 | 28 | 0 | 1 | Cuba | Caribbean (Travel) | 2 | 2III_D.1.1 |
| OQ821436 | 2019 | 9 | 29 | 0 | 1 | Cuba | Caribbean (Travel) | 2 | 2III_C.2 |
| OQ821437 | 2019 | 9 | 29 | 0 | 1 | Cuba | Caribbean (Travel) | 2 | 2III_C.2 |
| OQ821336 | 2019 | 10 | 2 | 0 | 1 | Cuba | Caribbean (Travel) | 1 | 1V_D.1.1 |
| OQ821337 | 2019 | 10 | 4 | 0 | 1 | Cuba | Caribbean (Travel) | 1 | 1V_D.1.1 |
| OQ821358 | 2019 | 10 | 8 | 0 | 1 | Dominican-Republic | Caribbean (Travel) | 1 | 1V_E.2 |
| OQ821438 | 2019 | 10 | 10 | 0 | 1 | Cuba | Caribbean (Travel) | 2 | 2III_C.2 |
| OQ821439 | 2019 | 10 | 13 | 0 | 1 | Cuba | Caribbean (Travel) | 2 | 2III_C.2 |
| OQ821440 | 2019 | 10 | 14 | 0 | 1 | Cuba | Caribbean (Travel) | 2 | 2III_C.2 |
| OQ821441 | 2019 | 10 | 14 | 0 | 1 | Cuba | Caribbean (Travel) | 2 | 2III_D.1.1 |
| OQ821359 | 2019 | 10 | 17 | 0 | 1 | Dominican-Republic | Caribbean (Travel) | 1 | 1V_E.2 |
| OQ821608 | 2019 | 10 | 19 | 0 | 1 | Jamaica | Caribbean (Travel) | 3 | 3III_B.3.1 |
| OQ821380 | 2019 | 10 | 23 | 0 | 1 | Venezuela | South-America (Travel) | 1 | 1V_D.1 |
| OQ821442 | 2019 | 10 | 28 | 0 | 1 | Cuba | Caribbean (Travel) | 2 | 2III_D.1.1 |
| OQ821609 | 2019 | 11 | 6 | 0 | 1 | Jamaica | Caribbean (Travel) | 3 | 3III_B.3.1 |
| OQ821484 | 2019 | 11 | 6 | 0 | 1 | Mexico | North-America (Travel) | 2 | 2III_D.1.2 |
| OQ821443 | 2019 | 11 | 9 | 0 | 1 | Cuba | Caribbean (Travel) | 2 | 2III_C.2 |
| OQ821444 | 2019 | 11 | 13 | 0 | 1 | Cuba | Caribbean (Travel) | 2 | 2III_C.2 |
| OQ821445 | 2019 | 11 | 13 | 0 | 1 | Cuba | Caribbean (Travel) | 2 | 2III_C.2 |
| OQ821446 | 2019 | 11 | 24 | 0 | 1 | Cuba | Caribbean (Travel) | 2 | 2III_C.2 |
| OQ821447 | 2019 | 11 | 25 | 0 | 1 | Cuba | Caribbean (Travel) | 2 | 2III_C.2 |
| OQ821366 | 2019 | 12 | 5 | 0 | 1 | Mexico | North-America (Travel) | 1 | 1V_D.1 |
| OQ821499 | 2019 | 12 | 10 | 1 | 0 | Florida (USA) | Miami-Dade | 2 | 2III_C.2 |
| OQ821448 | 2019 | 12 | 11 | 0 | 1 | Cuba | Caribbean (Travel) | 2 | 2III_C.2 |
| OQ821492 | 2019 | 12 | 13 | 0 | 1 | Nicaragua | Central-America (Travel) | 2 | 2III_D.1.2 |
| OQ821325 | 2020 | 1 | 9 | 0 | 1 | Bolivia | South-America (Travel) | 1 | 1V_D.2 |
| OQ821338 | 2020 | 1 | 10 | 0 | 1 | Cuba | Caribbean (Travel) | 1 | 1V_D.1.1 |
| OQ821449 | 2020 | 1 | 15 | 0 | 1 | Cuba | Caribbean (Travel) | 2 | 2III_C.2 |
| OQ821610 | 2020 | 1 | 17 | 0 | 1 | Jamaica | Caribbean (Travel) | 3 | 3III_B.3.1 |
| OQ821326 | 2020 | 3 | 15 | 0 | 1 | Bolivia | South-America (Travel) | 1 | 1V_D.2 |
| OQ821624 | 2020 | 3 | 19 | 0 | 1 | Paraguay | South-America (Travel) | 4 | 4II_B.1.1 |
| OM833056 | 2020 | 6 | 23 | 1 | 0 | Florida (USA) | Monroe | 1 | 1V_E.2 |
| OM833058 | 2020 | 6 | 27 | 1 | 0 | Florida (USA) | Monroe | 1 | 1V_E.2 |
| OM833057 | 2020 | 7 | 29 | 1 | 0 | Florida (USA) | Monroe | 1 | 1V_E.2 |
| OQ821611 | 2020 | 11 | 7 | 0 | 1 | Saint-Lucia | Caribbean (Travel) | 3 | 3III_B.3.1 |
| OQ821339 | 2020 | 11 | 25 | 0 | 1 | Cuba | Caribbean (Travel) | 1 | 1V_D.1.1 |
| OQ821365 | 2020 | 12 | 18 | 0 | 1 | Cuba | Caribbean (Travel) | 1 | 1V_D.1.1 |
| OQ821502 | 2021 | 8 | 12 | 0 | 1 | Colombia | South-America (Travel) | 3 | 3III_C.1 |
| OQ821340 | 2021 | 11 | 16 | 0 | 1 | Cuba | Caribbean (Travel) | 1 | 1V_E.2 |
| OQ821450 | 2021 | 12 | 22 | 0 | 1 | Cuba | Caribbean (Travel) | 2 | 2III_D.1.1 |
| OQ821451 | 2021 | 12 | 24 | 0 | 1 | Cuba | Caribbean (Travel) | 2 | 2III_D.1.1 |
| OQ821452 | 2022 | 1 | 30 | 0 | 1 | Cuba | Caribbean (Travel) | 2 | 2III_C.2 |
| OQ821453 | 2022 | 2 | 19 | 0 | 1 | Cuba | Caribbean (Travel) | 2 | 2III_D.1.1 |
| OQ821341 | 2022 | 4 | 24 | 0 | 1 | Cuba | Caribbean (Travel) | 1 | 1V_D.1.1 |
| OQ821342 | 2022 | 5 | 16 | 0 | 1 | Cuba | Caribbean (Travel) | 1 | 1V_D.1.1 |
| OQ821454 | 2022 | 6 | 13 | 0 | 1 | Cuba | Caribbean (Travel) | 2 | 2III_C.2 |
| OQ821455 | 2022 | 6 | 15 | 0 | 1 | Cuba | Caribbean (Travel) | 2 | 2III_C.2 |
| OQ821639 | 2022 | 6 | 15 | 0 | 1 | Cuba | Caribbean (Travel) | 4 | 4II_B.1.2 |
| OQ821343 | 2022 | 6 | 17 | 0 | 1 | Cuba | Caribbean (Travel) | 1 | 1V_D.1 |
| OQ821637 | 2022 | 6 | 19 | 0 | 1 | Cuba | Caribbean (Travel) | 4 | 4II_B.1.2 |
| OQ821456 | 2022 | 6 | 20 | 0 | 1 | Cuba | Caribbean (Travel) | 2 | 2III_D.1.1 |
| OQ821344 | 2022 | 6 | 23 | 0 | 1 | Cuba | Caribbean (Travel) | 1 | 1V_D.1.1 |
| OQ821642 | 2022 | 6 | 24 | 0 | 1 | Cuba | Caribbean (Travel) | 4 | 4II_B.1.2 |
| OQ821457 | 2022 | 6 | 28 | 0 | 1 | Cuba | Caribbean (Travel) | 2 | 2III_D.1.1 |
| OQ821458 | 2022 | 6 | 28 | 0 | 1 | Cuba | Caribbean (Travel) | 2 | 2III_D.1.1 |
| OQ821459 | 2022 | 6 | 28 | 0 | 1 | Cuba | Caribbean (Travel) | 2 | 2III_D.1.1 |
| OQ821329 | 2022 | 6 | 29 | 0 | 1 | Brazil | South-America (Travel) | 1 | 1V_E.1 |
| OQ821481 | 2022 | 7 | 1 | 0 | 1 | India | South-Asia (Travel) | 2 | 2II_F.1.1 |
| OQ821345 | 2022 | 7 | 2 | 0 | 1 | Cuba | Caribbean (Travel) | 1 | 1V_E.2 |
| OQ821460 | 2022 | 7 | 4 | 0 | 1 | Cuba | Caribbean (Travel) | 2 | 2III_C.2 |
| OQ821640 | 2022 | 7 | 5 | 0 | 1 | Cuba | Caribbean (Travel) | 4 | 4II_B.1.2 |
| OQ821461 | 2022 | 7 | 6 | 0 | 1 | Cuba | Caribbean (Travel) | 2 | 2III_C.2 |
| OQ821462 | 2022 | 7 | 6 | 0 | 1 | Cuba | Caribbean (Travel) | 2 | 2III_C.2 |
| OQ821504 | 2022 | 7 | 6 | 0 | 1 | Cuba | Caribbean (Travel) | 3 | 3III_B.3.2 |
| OQ821485 | 2022 | 7 | 6 | 0 | 1 | Mexico | North-America (Travel) | 2 | 2III_D.1.2 |
| OQ821641 | 2022 | 7 | 11 | 0 | 1 | Cuba | Caribbean (Travel) | 4 | 4II_B.1.2 |
| OQ445953 | 2022 | 7 | 13 | 1 | 0 | Florida (USA) | Miami-Dade | 3 | 3III_B.3.2 |
| OQ821346 | 2022 | 7 | 14 | 0 | 1 | Cuba | Caribbean (Travel) | 1 | 1V_D.1 |
| OQ821463 | 2022 | 7 | 14 | 0 | 1 | Cuba | Caribbean (Travel) | 2 | 2III_D.1.1 |
| OQ821643 | 2022 | 7 | 15 | 0 | 1 | Cuba | Caribbean (Travel) | 4 | 4II_B.1.2 |
| OQ821505 | 2022 | 7 | 17 | 0 | 1 | Cuba | Caribbean (Travel) | 3 | 3III_B.3.2 |
| OQ821638 | 2022 | 7 | 21 | 0 | 1 | Cuba | Caribbean (Travel) | 4 | 4II_B.1.2 |
| OQ821506 | 2022 | 7 | 23 | 0 | 1 | Cuba | Caribbean (Travel) | 3 | 3III_B.3.2 |
| OQ821464 | 2022 | 7 | 26 | 0 | 1 | Cuba | Caribbean (Travel) | 2 | 2III_D.1.1 |
| OQ821507 | 2022 | 7 | 28 | 0 | 1 | Cuba | Caribbean (Travel) | 3 | 3III_B.3.2 |
| OQ821508 | 2022 | 7 | 28 | 0 | 1 | Cuba | Caribbean (Travel) | 3 | 3III_B.3.2 |
| OQ445948 | 2022 | 7 | 29 | 1 | 0 | Florida (USA) | Miami-Dade | 3 | 3III_B.3.2 |
| OQ821347 | 2022 | 8 | 2 | 0 | 1 | Cuba | Caribbean (Travel) | 1 | 1V_D.1 |
| OQ821466 | 2022 | 8 | 5 | 0 | 1 | Cuba | Caribbean (Travel) | 2 | 2III_D.1.1 |
| OQ821509 | 2022 | 8 | 5 | 0 | 1 | Cuba | Caribbean (Travel) | 3 | 3III_B.3.2 |
| OQ445947 | 2022 | 8 | 5 | 1 | 0 | Florida (USA) | Miami-Dade | 3 | 3III_B.3.2 |
| OQ821510 | 2022 | 8 | 6 | 0 | 1 | Cuba | Caribbean (Travel) | 3 | 3III_B.3.2 |
| OQ821467 | 2022 | 8 | 8 | 0 | 1 | Cuba | Caribbean (Travel) | 2 | 2III_C.2 |
| OQ821613 | 2022 | 8 | 8 | 1 | 0 | Florida (USA) | Miami-Dade | 3 | 3III_B.3.2 |
| OQ821511 | 2022 | 8 | 9 | 0 | 1 | Cuba | Caribbean (Travel) | 3 | 3III_B.3.2 |
| OQ445918 | 2022 | 8 | 9 | 1 | 0 | Florida (USA) | Miami-Dade | 3 | 3III_B.3.2 |
| OQ445946 | 2022 | 8 | 9 | 1 | 0 | Florida (USA) | Miami-Dade | 3 | 3III_B.3.2 |
| OQ821512 | 2022 | 8 | 10 | 0 | 1 | Cuba | Caribbean (Travel) | 3 | 3III_B.3.2 |
| OQ821614 | 2022 | 8 | 11 | 1 | 0 | Florida (USA) | Miami-Dade | 3 | 3III_B.3.2 |
| OQ821513 | 2022 | 8 | 12 | 0 | 1 | Cuba | Caribbean (Travel) | 3 | 3III_B.3.2 |
| OQ821514 | 2022 | 8 | 13 | 0 | 1 | Cuba | Caribbean (Travel) | 3 | 3III_B.3.2 |
| OQ821515 | 2022 | 8 | 14 | 0 | 1 | Cuba | Caribbean (Travel) | 3 | 3III_B.3.2 |
| OQ821516 | 2022 | 8 | 15 | 0 | 1 | Cuba | Caribbean (Travel) | 3 | 3III_B.3.2 |
| OQ445919 | 2022 | 8 | 15 | 1 | 0 | Florida (USA) | Miami-Dade | 3 | 3III_B.3.2 |
| OQ821517 | 2022 | 8 | 16 | 0 | 1 | Cuba | Caribbean (Travel) | 3 | 3III_B.3.2 |
| OQ821615 | 2022 | 8 | 17 | 1 | 0 | Florida (USA) | Miami-Dade | 3 | 3III_B.3.2 |
| OQ821468 | 2022 | 8 | 18 | 0 | 1 | Cuba | Caribbean (Travel) | 2 | 2III_C.2 |
| OQ821518 | 2022 | 8 | 18 | 0 | 1 | Cuba | Caribbean (Travel) | 3 | 3III_B.3.2 |
| OQ821519 | 2022 | 8 | 18 | 0 | 1 | Cuba | Caribbean (Travel) | 3 | 3III_B.3.2 |
| OQ821520 | 2022 | 8 | 18 | 0 | 1 | Cuba | Caribbean (Travel) | 3 | 3III_B.3.2 |
| OQ821521 | 2022 | 8 | 18 | 0 | 1 | Cuba | Caribbean (Travel) | 3 | 3III_B.3.2 |
| OQ821522 | 2022 | 8 | 18 | 0 | 1 | Cuba | Caribbean (Travel) | 3 | 3III_B.3.2 |
| OQ821523 | 2022 | 8 | 18 | 0 | 1 | Cuba | Caribbean (Travel) | 3 | 3III_B.3.2 |
| OQ445920 | 2022 | 8 | 18 | 1 | 0 | Florida (USA) | Miami-Dade | 3 | 3III_B.3.2 |
| OQ821482 | 2022 | 8 | 19 | 0 | 1 | India | South-Asia (Travel) | 2 | 2II_F.1.1 |
| OQ821524 | 2022 | 8 | 20 | 0 | 1 | Cuba | Caribbean (Travel) | 3 | 3III_B.3.2 |
| OQ919693 | 2022 | 8 | 20 | 0 | 1 | Cuba | Caribbean (Travel) | 3 | 3III_B.3.2 |
| OQ821525 | 2022 | 8 | 21 | 0 | 1 | Cuba | Caribbean (Travel) | 3 | 3III_B.3.2 |
| OQ821526 | 2022 | 8 | 23 | 0 | 1 | Cuba | Caribbean (Travel) | 3 | 3III_B.3.2 |
| OQ821527 | 2022 | 8 | 23 | 0 | 1 | Cuba | Caribbean (Travel) | 3 | 3III_B.3.2 |
| OQ821528 | 2022 | 8 | 24 | 0 | 1 | Cuba | Caribbean (Travel) | 3 | 3III_B.3.2 |
| OQ821529 | 2022 | 8 | 24 | 0 | 1 | Cuba | Caribbean (Travel) | 3 | 3III_B.3.2 |
| OQ821530 | 2022 | 8 | 24 | 0 | 1 | Cuba | Caribbean (Travel) | 3 | 3III_B.3.2 |
| OQ821646 | 2022 | 8 | 24 | 0 | 1 | El-Salvador | Central-America (Travel) | 4 | 4II_B.2 |
| OQ821469 | 2022 | 8 | 26 | 0 | 1 | Cuba | Caribbean (Travel) | 2 | 2III_C.2 |
| OQ821348 | 2022 | 8 | 28 | 0 | 1 | Cuba | Caribbean (Travel) | 1 | 1V_D.1.1 |
| OQ821531 | 2022 | 8 | 28 | 0 | 1 | Cuba | Caribbean (Travel) | 3 | 3III_B.3.2 |
| OQ445921 | 2022 | 8 | 28 | 1 | 0 | Florida (USA) | Miami-Dade | 3 | 3III_B.3.2 |
| OQ445922 | 2022 | 8 | 29 | 1 | 0 | Florida (USA) | Miami-Dade | 3 | 3III_B.3.2 |
| OQ821532 | 2022 | 8 | 30 | 0 | 1 | Cuba | Caribbean (Travel) | 3 | 3III_B.3.2 |
| OQ821533 | 2022 | 8 | 31 | 0 | 1 | Cuba | Caribbean (Travel) | 3 | 3III_B.3.2 |
| OQ821534 | 2022 | 8 | 31 | 0 | 1 | Cuba | Caribbean (Travel) | 3 | 3III_B.3.2 |
| OQ821349 | 2022 | 9 | 1 | 0 | 1 | Cuba | Caribbean (Travel) | 1 | 1V_D.1.1 |
| OQ821535 | 2022 | 9 | 1 | 0 | 1 | Cuba | Caribbean (Travel) | 3 | 3III_B.3.2 |
| OQ821536 | 2022 | 9 | 3 | 0 | 1 | Cuba | Caribbean (Travel) | 3 | 3III_B.3.2 |
| OQ821473 | 2022 | 9 | 3 | 0 | 1 | Dominican-Republic | Caribbean (Travel) | 2 | 2III_C.1.2 |
| OQ821537 | 2022 | 9 | 5 | 0 | 1 | Cuba | Caribbean (Travel) | 3 | 3III_B.3.2 |
| OQ821538 | 2022 | 9 | 5 | 0 | 1 | Cuba | Caribbean (Travel) | 3 | 3III_B.3.2 |
| OQ445923 | 2022 | 9 | 5 | 1 | 0 | Florida (USA) | Miami-Dade | 3 | 3III_B.3.2 |
| OQ821539 | 2022 | 9 | 7 | 0 | 1 | Cuba | Caribbean (Travel) | 3 | 3III_B.3.2 |
| OQ821540 | 2022 | 9 | 8 | 0 | 1 | Cuba | Caribbean (Travel) | 3 | 3III_B.3.2 |
| OQ821645 | 2022 | 9 | 10 | 0 | 1 | Nicaragua | Central-America (Travel) | 4 | 4II_B.2 |
| OQ821541 | 2022 | 9 | 12 | 0 | 1 | Cuba | Caribbean (Travel) | 3 | 3III_B.3.2 |
| OQ821542 | 2022 | 9 | 13 | 0 | 1 | Cuba | Caribbean (Travel) | 3 | 3III_B.3.2 |
| OQ821543 | 2022 | 9 | 13 | 0 | 1 | Cuba | Caribbean (Travel) | 3 | 3III_B.3.2 |
| OQ821544 | 2022 | 9 | 14 | 0 | 1 | Cuba | Caribbean (Travel) | 3 | 3III_B.3.2 |
| OQ445925 | 2022 | 9 | 14 | 1 | 0 | Florida (USA) | Miami-Dade | 3 | 3III_B.3.2 |
| OQ821545 | 2022 | 9 | 15 | 0 | 1 | Cuba | Caribbean (Travel) | 3 | 3III_B.3.2 |
| OQ821546 | 2022 | 9 | 17 | 0 | 1 | Cuba | Caribbean (Travel) | 3 | 3III_B.3.2 |
| OQ821547 | 2022 | 9 | 17 | 0 | 1 | Cuba | Caribbean (Travel) | 3 | 3III_B.3.2 |
| OQ821350 | 2022 | 9 | 19 | 0 | 1 | Cuba | Caribbean (Travel) | 1 | 1V_D.1 |
| OQ821548 | 2022 | 9 | 21 | 0 | 1 | Cuba | Caribbean (Travel) | 3 | 3III_B.3.2 |
| OQ821549 | 2022 | 9 | 22 | 0 | 1 | Cuba | Caribbean (Travel) | 3 | 3III_B.3.2 |
| OQ821550 | 2022 | 9 | 22 | 0 | 1 | Cuba | Caribbean (Travel) | 3 | 3III_B.3.2 |
| OQ919689 | 2022 | 9 | 22 | 1 | 0 | Florida (USA) | Miami-Dade | 3 | 3III_B.3.2 |
| OQ821551 | 2022 | 9 | 23 | 0 | 1 | Cuba | Caribbean (Travel) | 3 | 3III_B.3.2 |
| OQ821552 | 2022 | 9 | 23 | 0 | 1 | Cuba | Caribbean (Travel) | 3 | 3III_B.3.2 |
| OQ821553 | 2022 | 9 | 23 | 0 | 1 | Cuba | Caribbean (Travel) | 3 | 3III_B.3.2 |
| OQ821554 | 2022 | 9 | 25 | 0 | 1 | Cuba | Caribbean (Travel) | 3 | 3III_B.3.2 |
| OQ821555 | 2022 | 9 | 25 | 0 | 1 | Cuba | Caribbean (Travel) | 3 | 3III_B.3.2 |
| OQ821556 | 2022 | 9 | 25 | 0 | 1 | Cuba | Caribbean (Travel) | 3 | 3III_B.3.2 |
| OQ445927 | 2022 | 9 | 25 | 1 | 0 | Florida (USA) | Miami-Dade | 3 | 3III_B.3.2 |
| OQ445929 | 2022 | 9 | 25 | 1 | 0 | Florida (USA) | Miami-Dade | 3 | 3III_B.3.2 |
| OQ821470 | 2022 | 9 | 26 | 0 | 1 | Cuba | Caribbean (Travel) | 2 | 2III_D.1.1 |
| OQ821557 | 2022 | 9 | 26 | 0 | 1 | Cuba | Caribbean (Travel) | 3 | 3III_B.3.2 |
| OQ821558 | 2022 | 9 | 26 | 0 | 1 | Cuba | Caribbean (Travel) | 3 | 3III_B.3.2 |
| OR150743 | 2022 | 9 | 26 | 1 | 0 | Florida (USA) | Miami-Dade | 2 | 2III_D.1.1 |
| OQ821559 | 2022 | 9 | 29 | 0 | 1 | Cuba | Caribbean (Travel) | 3 | 3III_B.3.2 |
| OQ821560 | 2022 | 9 | 29 | 0 | 1 | Cuba | Caribbean (Travel) | 3 | 3III_B.3.2 |
| OQ821561 | 2022 | 9 | 30 | 0 | 1 | Cuba | Caribbean (Travel) | 3 | 3III_B.3.2 |
| OQ821562 | 2022 | 10 | 2 | 0 | 1 | Cuba | Caribbean (Travel) | 3 | 3III_B.3.2 |
| OQ821616 | 2022 | 10 | 4 | 1 | 0 | Florida (USA) | Miami-Dade | 3 | 3III_B.3.2 |
| OQ821563 | 2022 | 10 | 5 | 0 | 1 | Cuba | Caribbean (Travel) | 3 | 3III_B.3.2 |
| OQ919690 | 2022 | 10 | 5 | 0 | 1 | Cuba | Caribbean (Travel) | 3 | 3III_B.3.2 |
| OQ821564 | 2022 | 10 | 7 | 0 | 1 | Cuba | Caribbean (Travel) | 3 | 3III_B.3.2 |
| OQ445930 | 2022 | 10 | 10 | 1 | 0 | Florida (USA) | Miami-Dade | 3 | 3III_B.3.2 |
| OQ821617 | 2022 | 10 | 10 | 1 | 0 | Florida (USA) | Miami-Dade | 3 | 3III_B.3.2 |
| OQ821565 | 2022 | 10 | 14 | 0 | 1 | Cuba | Caribbean (Travel) | 3 | 3III_B.3.2 |
| OQ821566 | 2022 | 10 | 15 | 0 | 1 | Cuba | Caribbean (Travel) | 3 | 3III_B.3.2 |
| OQ821618 | 2022 | 10 | 15 | 1 | 0 | Florida (USA) | Miami-Dade | 3 | 3III_B.3.2 |
| OQ821567 | 2022 | 10 | 19 | 0 | 1 | Cuba | Caribbean (Travel) | 3 | 3III_B.3.2 |
| OQ445931 | 2022 | 10 | 19 | 1 | 0 | Florida (USA) | Miami-Dade | 3 | 3III_B.3.2 |
| OQ821568 | 2022 | 10 | 20 | 0 | 1 | Cuba | Caribbean (Travel) | 3 | 3III_B.3.2 |
| OQ445934 | 2022 | 10 | 20 | 1 | 0 | Florida (USA) | Miami-Dade | 3 | 3III_B.3.2 |
| OQ821569 | 2022 | 10 | 21 | 0 | 1 | Cuba | Caribbean (Travel) | 3 | 3III_B.3.2 |
| OQ821644 | 2022 | 10 | 22 | 0 | 1 | Nicaragua | Central-America (Travel) | 4 | 4II_B.2 |
| OQ445932 | 2022 | 10 | 23 | 1 | 0 | Florida (USA) | Miami-Dade | 3 | 3III_B.3.2 |
| OQ445933 | 2022 | 10 | 24 | 1 | 0 | Florida (USA) | Miami-Dade | 3 | 3III_B.3.2 |
| OQ445935 | 2022 | 10 | 26 | 1 | 0 | Florida (USA) | Miami-Dade | 3 | 3III_B.3.2 |
| OQ821570 | 2022 | 10 | 30 | 0 | 1 | Cuba | Caribbean (Travel) | 3 | 3III_B.3.2 |
| OQ821571 | 2022 | 11 | 3 | 0 | 1 | Cuba | Caribbean (Travel) | 3 | 3III_B.3.2 |
| OQ821572 | 2022 | 11 | 3 | 0 | 1 | Cuba | Caribbean (Travel) | 3 | 3III_B.3.2 |
| OQ445967 | 2022 | 11 | 3 | 1 | 0 | Florida (USA) | Miami-Dade | 4 | 4II_B.2 |
| OQ821573 | 2022 | 11 | 4 | 0 | 1 | Cuba | Caribbean (Travel) | 3 | 3III_B.3.2 |
| OQ821574 | 2022 | 11 | 7 | 0 | 1 | Cuba | Caribbean (Travel) | 3 | 3III_B.3.2 |
| OQ919691 | 2022 | 11 | 8 | 1 | 0 | Florida (USA) | Miami-Dade | 3 | 3III_B.3.2 |
| OQ821575 | 2022 | 11 | 10 | 0 | 1 | Cuba | Caribbean (Travel) | 3 | 3III_B.3.2 |
| OQ821576 | 2022 | 11 | 14 | 0 | 1 | Cuba | Caribbean (Travel) | 3 | 3III_B.3.2 |
| OQ821636 | 2022 | 11 | 15 | 0 | 1 | Cuba | Caribbean (Travel) | 4 | 4II_B.1.2 |
| OR162320 | 2022 | 11 | 15 | 1 | 0 | Florida (USA) | Miami-Dade | 4 | 4II_B.1.2 |
| OQ821619 | 2022 | 11 | 17 | 1 | 0 | Florida (USA) | Miami-Dade | 3 | 3III_B.3.2 |
| OQ821577 | 2022 | 11 | 18 | 0 | 1 | Cuba | Caribbean (Travel) | 3 | 3III_B.3.2 |
| OQ821578 | 2022 | 11 | 18 | 0 | 1 | Cuba | Caribbean (Travel) | 3 | 3III_B.3.2 |
| OQ821579 | 2022 | 11 | 20 | 0 | 1 | Cuba | Caribbean (Travel) | 3 | 3III_B.3.2 |
| OQ821620 | 2022 | 11 | 21 | 1 | 0 | Florida (USA) | Miami-Dade | 3 | 3III_B.3.2 |
| OQ821580 | 2022 | 11 | 25 | 0 | 1 | Cuba | Caribbean (Travel) | 3 | 3III_B.3.2 |
| PP709312 | 2022 | 11 | 25 | 1 | 0 | Florida (USA) | Miami-Dade | 3 | 3III_B.3.2 |
| OR162311 | 2022 | 11 | 27 | 1 | 0 | Florida (USA) | Miami-Dade | 3 | 3III_B.3.2 |
| OR162313 | 2022 | 11 | 27 | 1 | 0 | Florida (USA) | Miami-Dade | 3 | 3III_B.3.2 |
| OQ821621 | 2022 | 11 | 28 | 1 | 0 | Florida (USA) | Miami-Dade | 3 | 3III_B.3.2 |
| OQ821581 | 2022 | 11 | 29 | 0 | 1 | Cuba | Caribbean (Travel) | 3 | 3III_B.3.2 |
| OQ821471 | 2022 | 11 | 30 | 0 | 1 | Cuba | Caribbean (Travel) | 2 | 2III_D.1.1 |
| OQ821582 | 2022 | 11 | 30 | 0 | 1 | Cuba | Caribbean (Travel) | 3 | 3III_B.3.2 |
| OQ821583 | 2022 | 11 | 30 | 0 | 1 | Cuba | Caribbean (Travel) | 3 | 3III_B.3.2 |
| OQ821584 | 2022 | 11 | 30 | 0 | 1 | Cuba | Caribbean (Travel) | 3 | 3III_B.3.2 |
| OQ821585 | 2022 | 12 | 2 | 0 | 1 | Cuba | Caribbean (Travel) | 3 | 3III_B.3.2 |
| OR162312 | 2022 | 12 | 2 | 1 | 0 | Florida (USA) | Miami-Dade | 3 | 3III_B.3.2 |
| OR162316 | 2022 | 12 | 2 | 1 | 0 | Florida (USA) | Miami-Dade | 3 | 3III_B.3.2 |
| OQ821351 | 2022 | 12 | 3 | 0 | 1 | Cuba | Caribbean (Travel) | 1 | 1V_D.1.1 |
| OQ821586 | 2022 | 12 | 4 | 0 | 1 | Cuba | Caribbean (Travel) | 3 | 3III_B.3.2 |
| OQ821587 | 2022 | 12 | 4 | 0 | 1 | Cuba | Caribbean (Travel) | 3 | 3III_B.3.2 |
| OQ919692 | 2022 | 12 | 4 | 0 | 1 | Cuba | Caribbean (Travel) | 3 | 3III_B.3.2 |
| OR162314 | 2022 | 12 | 4 | 1 | 0 | Florida (USA) | Miami-Dade | 3 | 3III_B.3.2 |
| OR162317 | 2022 | 12 | 4 | 1 | 0 | Florida (USA) | Miami-Dade | 3 | 3III_B.3.2 |
| OQ821588 | 2022 | 12 | 6 | 0 | 1 | Cuba | Caribbean (Travel) | 3 | 3III_B.3.2 |
| OQ821589 | 2022 | 12 | 6 | 0 | 1 | Cuba | Caribbean (Travel) | 3 | 3III_B.3.2 |
| OR162315 | 2022 | 12 | 6 | 1 | 0 | Florida (USA) | Miami-Dade | 3 | 3III_B.3.2 |
| OQ821352 | 2022 | 12 | 7 | 0 | 1 | Cuba | Caribbean (Travel) | 1 | 1V_D.1.1 |
| OQ821367 | 2022 | 12 | 7 | 0 | 1 | Mexico | North-America (Travel) | 1 | 1V_D.1.2 |
| OQ821590 | 2022 | 12 | 8 | 0 | 1 | Cuba | Caribbean (Travel) | 3 | 3III_B.3.2 |
| OR162318 | 2022 | 12 | 10 | 1 | 0 | Florida (USA) | Miami-Dade | 3 | 3III_B.3.2 |
| OQ821622 | 2022 | 12 | 11 | 1 | 0 | Florida (USA) | Miami-Dade | 3 | 3III_B.3.2 |
| OQ821591 | 2022 | 12 | 15 | 0 | 1 | Cuba | Caribbean (Travel) | 3 | 3III_B.3.2 |
| OQ821592 | 2022 | 12 | 16 | 0 | 1 | Cuba | Caribbean (Travel) | 3 | 3III_B.3.2 |
| OQ821593 | 2022 | 12 | 16 | 0 | 1 | Cuba | Caribbean (Travel) | 3 | 3III_B.3.2 |
| OQ821594 | 2022 | 12 | 18 | 0 | 1 | Cuba | Caribbean (Travel) | 3 | 3III_B.3.2 |
| OQ821595 | 2022 | 12 | 18 | 0 | 1 | Cuba | Caribbean (Travel) | 3 | 3III_B.3.2 |
| OQ821596 | 2022 | 12 | 18 | 0 | 1 | Cuba | Caribbean (Travel) | 3 | 3III_B.3.2 |
| OR150746 | 2022 | 12 | 18 | 1 | 0 | Florida (USA) | Miami-Dade | 3 | 3III_B.3.2 |
| OR162319 | 2022 | 12 | 18 | 1 | 0 | Florida (USA) | Miami-Dade | 3 | 3III_B.3.2 |
| OR150744 | 2022 | 12 | 19 | 1 | 0 | Florida (USA) | Miami-Dade | 3 | 3III_B.3.2 |
| OQ821635 | 2022 | 12 | 20 | 0 | 1 | Cuba | Caribbean (Travel) | 4 | 4II_B.1.2 |
| OQ821597 | 2022 | 12 | 21 | 0 | 1 | Cuba | Caribbean (Travel) | 3 | 3III_B.3.2 |
| OQ821598 | 2022 | 12 | 22 | 0 | 1 | Cuba | Caribbean (Travel) | 3 | 3III_B.3.2 |
| OQ821599 | 2022 | 12 | 22 | 0 | 1 | Cuba | Caribbean (Travel) | 3 | 3III_B.3.2 |
| OR150745 | 2022 | 12 | 22 | 1 | 0 | Florida (USA) | Miami-Dade | 3 | 3III_B.3.2 |
| OQ821623 | 2022 | 12 | 23 | 1 | 0 | Florida (USA) | Miami-Dade | 3 | 3III_B.3.2 |
| OQ821600 | 2023 | 1 | 1 | 0 | 1 | Cuba | Caribbean (Travel) | 3 | 3III_B.3.2 |
| OR150748 | 2023 | 1 | 1 | 1 | 0 | Florida (USA) | Polk | 3 | 3III_B.3.2 |
| OR150749 | 2023 | 1 | 6 | 1 | 0 | Florida (USA) | Orange | 3 | 3III_B.3.2 |
| OR654284 | 2023 | 7 | 3 | 1 | 0 | Florida (USA) | Miami-Dade | 2 | 2II_F.1.1.2 |
| OR771126 | 2023 | 7 | 12 | 1 | 0 | Florida (USA) | Miami-Dade | 3 | 3III_B.3.2 |
| OR771131 | 2023 | 7 | 26 | 1 | 0 | Florida (USA) | Miami-Dade | 3 | 3III_B.3.2 |
| OR771136 | 2023 | 7 | 27 | 1 | 0 | Florida (USA) | Miami-Dade | 3 | 3III_B.3.2 |
| OR771144 | 2023 | 8 | 4 | 1 | 0 | Florida (USA) | Miami-Dade | 3 | 3III_B.3.2 |
| OR771158 | 2023 | 8 | 22 | 1 | 0 | Florida (USA) | Miami-Dade | 3 | 3III_B.3.2 |
| OR771167 | 2023 | 8 | 31 | 1 | 0 | Florida (USA) | Miami-Dade | 3 | 3III_B.3.2 |
| OR771177 | 2023 | 9 | 6 | 1 | 0 | Florida (USA) | Miami-Dade | 3 | 3III_B.3.2 |
| OR771179 | 2023 | 9 | 11 | 1 | 0 | Florida (USA) | Miami-Dade | 3 | 3III_B.3.2 |
| OR771180 | 2023 | 9 | 11 | 1 | 0 | Florida (USA) | Miami-Dade | 3 | 3III_B.3.2 |
| OR771183 | 2023 | 9 | 12 | 1 | 0 | Florida (USA) | Miami-Dade | 2 | 2II_F.1.1.2 |
| OR771181 | 2023 | 9 | 12 | 1 | 0 | Florida (USA) | Miami-Dade | 3 | 3III_B.3.2 |
| OR771185 | 2023 | 9 | 16 | 1 | 0 | Florida (USA) | Miami-Dade | 3 | 3III_B.3.2 |
| OR821940 | 2023 | 9 | 19 | 1 | 0 | Florida (USA) | Miami-Dade | 3 | 3III_B.3.2 |
| OR771182 | 2023 | 9 | 20 | 1 | 0 | Florida (USA) | Miami-Dade | 3 | 3III_B.3.2 |
| OR821936 | 2023 | 9 | 21 | 1 | 0 | Florida (USA) | Miami-Dade | 3 | 3III_B.3.2 |
| OR771188 | 2023 | 9 | 26 | 1 | 0 | Florida (USA) | Miami-Dade | 2 | 2II_F.1.1.2 |
| OR771189 | 2023 | 9 | 26 | 1 | 0 | Florida (USA) | Miami-Dade | 2 | 2II_F.1.1 |
| OR821944 | 2023 | 9 | 26 | 1 | 0 | Florida (USA) | Miami-Dade | 2 | 2II_F.1.1.2 |
| OR771195 | 2023 | 9 | 26 | 1 | 0 | Florida (USA) | Miami-Dade | 3 | 3III_B.3.2 |
| OR821941 | 2023 | 9 | 26 | 1 | 0 | Florida (USA) | Miami-Dade | 3 | 3III_B.3.2 |
| OR821942 | 2023 | 9 | 27 | 1 | 0 | Florida (USA) | Miami-Dade | 3 | 3III_B.3.2 |
| OR821946 | 2023 | 9 | 28 | 1 | 0 | Florida (USA) | Miami-Dade | 2 | 2II_F.1.1.2 |
| OR821945 | 2023 | 9 | 28 | 1 | 0 | Florida (USA) | Miami-Dade | 3 | 3III_B.3.2 |
| OR771190 | 2023 | 9 | 29 | 1 | 0 | Florida (USA) | Miami-Dade | 3 | 3III_B.3.2 |
| OR771193 | 2023 | 9 | 30 | 1 | 0 | Florida (USA) | Miami-Dade | 2 | 2II_F.1.1 |
| OR771191 | 2023 | 9 | 30 | 1 | 0 | Florida (USA) | Miami-Dade | 3 | 3III_B.3.2 |
| OR821948 | 2023 | 9 | 30 | 1 | 0 | Florida (USA) | Miami-Dade | 3 | 3III_B.3.2 |
| OR821950 | 2023 | 10 | 2 | 1 | 0 | Florida (USA) | Miami-Dade | 3 | 3III_B.3.2 |
| OR771199 | 2023 | 10 | 3 | 1 | 0 | Florida (USA) | Miami-Dade | 3 | 3III_B.3.2 |
| OR771192 | 2023 | 10 | 4 | 1 | 0 | Florida (USA) | Miami-Dade | 3 | 3III_B.3.2 |
| OR821952 | 2023 | 10 | 5 | 1 | 0 | Florida (USA) | Miami-Dade | 3 | 3III_B.3.2 |
| OR821949 | 2023 | 10 | 6 | 1 | 0 | Florida (USA) | Miami-Dade | 3 | 3III_B.3.2 |
| OR821951 | 2023 | 10 | 7 | 1 | 0 | Florida (USA) | Miami-Dade | 3 | 3III_B.3.2 |
| OR821955 | 2023 | 10 | 9 | 1 | 0 | Florida (USA) | Miami-Dade | 3 | 3III_B.3.2 |
| OR821954 | 2023 | 10 | 10 | 1 | 0 | Florida (USA) | Miami-Dade | 3 | 3III_B.3.2 |
| OR821958 | 2023 | 10 | 10 | 1 | 0 | Florida (USA) | Miami-Dade | 3 | 3III_B.3.2 |
| OR821962 | 2023 | 10 | 11 | 1 | 0 | Florida (USA) | Miami-Dade | 2 | 2II_F.1.1.5 |
| OR977072 | 2023 | 10 | 13 | 1 | 0 | Florida (USA) | Miami-Dade | 3 | 3III_B.3.2 |
| OR821964 | 2023 | 10 | 14 | 1 | 0 | Florida (USA) | Miami-Dade | 3 | 3III_B.3.2 |
| OR977076 | 2023 | 10 | 17 | 1 | 0 | Florida (USA) | Miami-Dade | 3 | 3III_B.3.2 |
| OR977080 | 2023 | 10 | 20 | 1 | 0 | Florida (USA) | Miami-Dade | 2 | 2II_F.1.1 |
| OR977077 | 2023 | 10 | 22 | 1 | 0 | Florida (USA) | Miami-Dade | 3 | 3III_B.3.2 |
| OR977081 | 2023 | 10 | 25 | 1 | 0 | Florida (USA) | Miami-Dade | 3 | 3III_B.3.2 |
| OR977085 | 2023 | 10 | 28 | 1 | 0 | Florida (USA) | Miami-Dade | 3 | 3III_B.3.2 |
| OR977086 | 2023 | 11 | 1 | 1 | 0 | Florida (USA) | Miami-Dade | 3 | 3III_B.3.2 |
| OR977088 | 2023 | 11 | 7 | 1 | 0 | Florida (USA) | Miami-Dade | 3 | 3III_B.3.2 |
| PP692455 | 2023 | 11 | 12 | 1 | 0 | Florida (USA) | Miami-Dade | 3 | 3III_B.3.2 |
| OR977091 | 2023 | 11 | 15 | 1 | 0 | Florida (USA) | Miami-Dade | 3 | 3III_B.3.2 |
| OR977092 | 2023 | 11 | 15 | 1 | 0 | Florida (USA) | Miami-Dade | 3 | 3III_B.3.2 |
| OR977099 | 2023 | 11 | 19 | 1 | 0 | Florida (USA) | Miami-Dade | 3 | 3III_B.3.2 |
| PP709309 | 2023 | 11 | 20 | 1 | 0 | Florida (USA) | Hardee | 1 | 1V_D.1 |
| OR977097 | 2023 | 11 | 20 | 1 | 0 | Florida (USA) | Miami-Dade | 3 | 3III_B.3.2 |
| PP692465 | 2023 | 12 | 12 | 1 | 0 | Florida (USA) | Miami-Dade | 3 | 3III_B.3.2 |
| PP708029 | 2024 | 1 | 2 | 1 | 0 | Florida (USA) | Miami-Dade | 3 | 3III_B.3.2 |
| PP708030 | 2024 | 1 | 8 | 1 | 0 | Florida (USA) | Miami-Dade | 3 | 3III_B.3.2 |
| PP708038 | 2024 | 1 | 20 | 1 | 0 | Florida (USA) | Miami-Dade | 3 | 3III_B.3.2 |
| PQ340822 | 2024 | 6 | 25 | 1 | 0 | Florida (USA) | Miami-Dade | 3 | 3III_B.3.2 |
| PQ527045 | 2024 | 8 | 13 | 1 | 0 | Florida (USA) | Hillsborough | 3 | 3III_B.3.2 |
| PQ617129 | 2024 | 8 | 26 | 1 | 0 | Florida (USA) | Pasco | 3 | 3III_B.3.2 |
| PQ617146 | 2024 | 9 | 8 | 1 | 0 | Florida (USA) | Miami-Dade | 3 | 3III_B.3.2 |
| PQ617120 | 2024 | 9 | 25 | 1 | 0 | Florida (USA) | Palm-Beach | 3 | 3III_B.3.2 |

**Table S3.** Transmission clusters of local DENV cases across counties in Florida. . This includes information on the county in which the cases were reported (sample region), major lineage, cluster size, inferred introduction sources and time, and the earliest and latest sampling dates of local dengue cases.

| Transmission cluster | Sample region (DTA) | Major lineage | Cluster size | Source region (DTA) | Posterior probability of source region | Median introduction time | Introduction time 95% credible interval | Earliest local sample date | Latest local sample date |
| --- | --- | --- | --- | --- | --- | --- | --- | --- | --- |
| Cluster 1 | Monroe | 1V_C | 8 | Central-America | 1 | 2010.1 | (2009.35,2010.44) | 2010.48 | 2010.88 |
| Cluster 2 | Miami-Dade | 2II_F | 4 | South-America | 1 | 2023.48 | (2023.28,2023.62) | 2023.7 | 2023.74 |
| Cluster 3 | Miami-Dade | 3III_B | 4 | Caribbean | 0.85 | 2021.95 | (2021.71,2022.13) | 2022.65 | 2022.81 |
| Cluster 4 | Miami-Dade | 3III_B | 27 | Caribbean | 1 | 2023.27 | (2023.09,2023.45) | 2023.64 | 2024.05 |
| Cluster 5 | Miami-Dade | 3III_B | 12 | Caribbean | 0.94 | 2023.31 | (2023.09,2023.54) | 2023.69 | 2024.02 |
| Cluster 6 | Miami-Dade | 3III_B | 6 | Caribbean | 0.72 | 2022.57 | (2022.37,2022.71) | 2022.72 | 2022.91 |

**Table S4.** Genomic sequences sampled from locally-acquired dengue infections in Florida from 2010 to 2024 and their inferred introduction sources and times from DTA analysis. This includes their GenBank accession numbers, sample collection date, assigned major lineage, trait used in DTA analysis (sample region (DTA)). The following columns are populated from our DTA analysis, including their inferred trait (source region (DTA)), posterior probability of inferred trait, transmission cluster membership, and introduction time (median height of transition node and its corresponding 95% credible interval). Introduction times can only be inferred for when a transition node has two or more descending tips belonging to a local dengue sample. Transmission cluster membership is defined by transition nodes with three or more descending tips belonging to a local dengue sample.

| GenBank Accession | Collection date | Collection date (decimal date) | Major lineage | Sample region (DTA) | Source region (DTA) | Posterior probability of source region | Transmission cluster | Median introduction time | Introduction time 95% credible interval |
| --- | --- | --- | --- | --- | --- | --- | --- | --- | --- |
| OQ821370 | 6/24/10 | 2010.48 | 1V_C | Monroe | Central-America | 1 | Cluster 1 | 2010.1 | (2009.35,2010.44) |
| OQ821371 | 6/30/10 | 2010.49 | 1V_C | Monroe | Central-America | 1 | Cluster 1 | 2010.1 | (2009.35,2010.44) |
| OQ821372 | 8/6/10 | 2010.59 | 1V_C | Monroe | Central-America | 1 | Cluster 1 | 2010.1 | (2009.35,2010.44) |
| OQ821373 | 8/11/10 | 2010.61 | 1V_C | Monroe | Central-America | 1 | Cluster 1 | 2010.1 | (2009.35,2010.44) |
| OQ821374 | 8/27/10 | 2010.65 | 1V_C | Monroe | Central-America | 1 | Cluster 1 | 2010.1 | (2009.35,2010.44) |
| JQ675358 | 2010-10 | 2010.75 | 1V_C | Monroe | Central-America | 1 | Cluster 1 | 2010.1 | (2009.35,2010.44) |
| OQ821375 | 10/11/10 | 2010.78 | 1V_C | Monroe | Central-America | 1 | Cluster 1 | 2010.1 | (2009.35,2010.44) |
| OQ821376 | 11/20/10 | 2010.88 | 1V_C | Monroe | Central-America | 1 | Cluster 1 | 2010.1 | (2009.35,2010.44) |
| OM833059 | 7/24/13 | 2013.56 | 1V_E | Monroe | Caribbean | 1 |  |  |  |
| OQ821377 | 8/29/13 | 2013.66 | 1V_E | Martin | Caribbean | 0.9 |  |  |  |
| OQ821612 | 7/2/14 | 2014.5 | 3I_A | Miami-Dade | Global | 1 |  |  |  |
| MN192436 | 2016 | 2016 | 4II_B | Manatee | Caribbean | 0.99 |  |  |  |
| KX702404 | 5/22/16 | 2016.39 | 2III_C | Alachua | South-America | 1 |  |  |  |
| OQ821496 | 7/11/19 | 2019.52 | 2III_D | Miami-Dade | Caribbean | 1 |  |  |  |
| OQ821497 | 8/29/19 | 2019.66 | 2III_D | Miami-Dade | Central-America | 1 |  |  |  |
| OQ821498 | 9/14/19 | 2019.7 | 2III_D | Miami-Dade | Central-America | 0.97 |  |  |  |
| OQ821378 | 9/20/19 | 2019.72 | 1III_A | Miami-Dade | Global | 1 |  |  |  |
| OQ821499 | 12/10/19 | 2019.94 | 2III_C | Miami-Dade | Caribbean | 1 |  |  |  |
| OM833056 | 6/23/20 | 2020.48 | 1V_E | Monroe | Caribbean | 0.98 |  | 2020.04 | (2019.25,2020.36) |
| OM833058 | 6/27/20 | 2020.49 | 1V_E | Monroe | Caribbean | 0.95 |  |  |  |
| OM833057 | 7/29/20 | 2020.57 | 1V_E | Monroe | Caribbean | 0.98 |  | 2020.04 | (2019.25,2020.36) |
| OQ445953 | 7/13/22 | 2022.53 | 3III_B | Miami-Dade | Caribbean | 1 |  |  |  |
| OQ445948 | 7/29/22 | 2022.57 | 3III_B | Miami-Dade | Caribbean | 1 |  | 2022.31 | (2021.99,2022.51) |
| OQ445947 | 8/5/22 | 2022.59 | 3III_B | Miami-Dade | Caribbean | 1 |  |  |  |
| OQ445918 | 8/9/22 | 2022.6 | 3III_B | Miami-Dade | Caribbean | 0.96 |  | 2021.86 | (2021.29,2022.25) |
| OQ445946 | 8/9/22 | 2022.6 | 3III_B | Miami-Dade | Caribbean | 1 |  | 2022.31 | (2021.99,2022.51) |
| OQ821613 | 8/8/22 | 2022.6 | 3III_B | Miami-Dade | Caribbean | 0.99 |  |  |  |
| OQ821614 | 8/11/22 | 2022.61 | 3III_B | Miami-Dade | Caribbean | 0.88 |  |  |  |
| OQ445919 | 8/15/22 | 2022.62 | 3III_B | Miami-Dade | Caribbean | 0.96 |  | 2021.86 | (2021.29,2022.25) |
| OQ821615 | 8/17/22 | 2022.62 | 3III_B | Miami-Dade | Caribbean | 1 |  |  |  |
| OQ445920 | 8/18/22 | 2022.63 | 3III_B | Miami-Dade | Caribbean | 0.93 |  |  |  |
| OQ445921 | 8/28/22 | 2022.65 | 3III_B | Miami-Dade | Caribbean | 0.85 | Cluster 3 | 2021.95 | (2021.71,2022.13) |
| OQ445922 | 8/29/22 | 2022.66 | 3III_B | Miami-Dade | Caribbean | 0.94 |  |  |  |
| OQ445923 | 9/5/22 | 2022.68 | 3III_B | Miami-Dade | Caribbean | 0.95 |  |  |  |
| OQ445925 | 9/14/22 | 2022.7 | 3III_B | Miami-Dade | Caribbean | 1 |  |  |  |
| OQ919689 | 9/22/22 | 2022.72 | 3III_B | Miami-Dade | Caribbean | 0.72 | Cluster 6 | 2022.57 | (2022.37,2022.71) |
| OR150743 | 9/26/22 | 2022.73 | 2III_D | Miami-Dade | Caribbean | 1 |  |  |  |
| OQ445927 | 9/25/22 | 2022.73 | 3III_B | Miami-Dade | Caribbean | 0.85 |  |  |  |
| OQ445929 | 9/25/22 | 2022.73 | 3III_B | Miami-Dade | Caribbean | 0.89 |  |  |  |
| OQ821616 | 10/4/22 | 2022.76 | 3III_B | Miami-Dade | Caribbean | 0.72 | Cluster 6 | 2022.57 | (2022.37,2022.71) |
| OQ445930 | 10/10/22 | 2022.77 | 3III_B | Miami-Dade | Caribbean | 0.85 | Cluster 3 | 2021.95 | (2021.71,2022.13) |
| OQ821617 | 10/10/22 | 2022.77 | 3III_B | Miami-Dade | Caribbean | 1 |  |  |  |
| OQ821618 | 10/15/22 | 2022.79 | 3III_B | Miami-Dade | Caribbean | 0.72 | Cluster 6 | 2022.57 | (2022.37,2022.71) |
| OQ445931 | 10/19/22 | 2022.8 | 3III_B | Miami-Dade | Caribbean | 0.85 | Cluster 3 | 2021.95 | (2021.71,2022.13) |
| OQ445934 | 10/20/22 | 2022.8 | 3III_B | Miami-Dade | Caribbean | 0.93 |  | 2022.2 | (2021.89,2022.57) |
| OQ445932 | 10/23/22 | 2022.81 | 3III_B | Miami-Dade | Caribbean | 0.95 |  |  |  |
| OQ445933 | 10/24/22 | 2022.81 | 3III_B | Miami-Dade | Caribbean | 0.85 | Cluster 3 | 2021.95 | (2021.71,2022.13) |
| OQ445935 | 10/26/22 | 2022.82 | 3III_B | Miami-Dade | Caribbean | 0.93 |  | 2022.2 | (2021.89,2022.57) |
| OQ445967 | 11/3/22 | 2022.84 | 4II_B | Miami-Dade | North-America | 0.82 |  |  |  |
| OQ919691 | 11/8/22 | 2022.85 | 3III_B | Miami-Dade | Caribbean | 1 |  | 2022.81 | (2022.65,2022.85) |
| OR162320 | 11/15/22 | 2022.87 | 4II_B | Miami-Dade | Caribbean | 0.67 |  |  |  |
| OQ821619 | 11/17/22 | 2022.88 | 3III_B | Miami-Dade | Caribbean | 0.72 | Cluster 6 | 2022.57 | (2022.37,2022.71) |
| OQ821620 | 11/21/22 | 2022.89 | 3III_B | Miami-Dade | Caribbean | 0.72 | Cluster 6 | 2022.57 | (2022.37,2022.71) |
| OR162311 | 11/27/22 | 2022.9 | 3III_B | Miami-Dade | Caribbean | 0.73 |  |  |  |
| OR162313 | 11/27/22 | 2022.9 | 3III_B | Miami-Dade | Caribbean | 1 |  | 2022.71 | (2022.37,2022.89) |
| PP709312 | 11/25/22 | 2022.9 | 3III_B | Miami-Dade | Caribbean | 0.7 |  |  |  |
| OQ821621 | 11/28/22 | 2022.91 | 3III_B | Miami-Dade | Caribbean | 0.72 | Cluster 6 | 2022.57 | (2022.37,2022.71) |
| OR162312 | 12/2/22 | 2022.92 | 3III_B | Miami-Dade | Caribbean | 0.6 |  |  |  |
| OR162314 | 12/4/22 | 2022.92 | 3III_B | Miami-Dade | Caribbean | 1 |  |  |  |
| OR162316 | 12/2/22 | 2022.92 | 3III_B | Miami-Dade | Caribbean | 1 |  |  |  |
| OR162317 | 12/4/22 | 2022.92 | 3III_B | Miami-Dade | Caribbean | 1 |  |  |  |
| OR162315 | 12/6/22 | 2022.93 | 3III_B | Miami-Dade | Caribbean | 1 |  |  |  |
| OQ821622 | 12/11/22 | 2022.94 | 3III_B | Miami-Dade | Caribbean | 1 |  | 2022.81 | (2022.65,2022.85) |
| OR162318 | 12/10/22 | 2022.94 | 3III_B | Miami-Dade | Caribbean | 1 |  | 2022.71 | (2022.37,2022.89) |
| OR150744 | 12/19/22 | 2022.96 | 3III_B | Miami-Dade | Caribbean | 1 |  |  |  |
| OR150746 | 12/18/22 | 2022.96 | 3III_B | Miami-Dade | Caribbean | 1 |  |  |  |
| OR162319 | 12/18/22 | 2022.96 | 3III_B | Miami-Dade | Caribbean | 1 |  |  |  |
| OR150745 | 12/22/22 | 2022.97 | 3III_B | Miami-Dade | Caribbean | 1 |  |  |  |
| OQ821623 | 12/23/22 | 2022.98 | 3III_B | Miami-Dade | Caribbean | 1 |  |  |  |
| OR150748 | 1/1/23 | 2023 | 3III_B | Polk | Caribbean | 1 |  |  |  |
| OR150749 | 1/6/23 | 2023.01 | 3III_B | Orange | Caribbean | 0.99 |  |  |  |
| OR654284 | 7/3/23 | 2023.5 | 2II_F | Miami-Dade | South-America | 1 |  |  |  |
| OR771126 | 7/12/23 | 2023.53 | 3III_B | Miami-Dade | Caribbean | 0.89 |  |  |  |
| OR771131 | 7/26/23 | 2023.56 | 3III_B | Miami-Dade | Caribbean | 0.94 |  |  |  |
| OR771136 | 7/27/23 | 2023.57 | 3III_B | Miami-Dade | Caribbean | 1 |  |  |  |
| OR771144 | 8/4/23 | 2023.59 | 3III_B | Miami-Dade | Caribbean | 1 |  |  |  |
| OR771158 | 8/22/23 | 2023.64 | 3III_B | Miami-Dade | Caribbean | 1 | Cluster 4 | 2023.27 | (2023.09,2023.45) |
| OR771167 | 8/31/23 | 2023.66 | 3III_B | Miami-Dade | Caribbean | 1 | Cluster 4 | 2023.27 | (2023.09,2023.45) |
| OR771177 | 9/6/23 | 2023.68 | 3III_B | Miami-Dade | Caribbean | 1 | Cluster 4 | 2023.27 | (2023.09,2023.45) |
| OR771179 | 9/11/23 | 2023.69 | 3III_B | Miami-Dade | Caribbean | 0.94 | Cluster 5 | 2023.31 | (2023.09,2023.54) |
| OR771180 | 9/11/23 | 2023.69 | 3III_B | Miami-Dade | Caribbean | 1 | Cluster 4 | 2023.27 | (2023.09,2023.45) |
| OR771183 | 9/12/23 | 2023.7 | 2II_F | Miami-Dade | South-America | 1 | Cluster 2 | 2023.48 | (2023.28,2023.62) |
| OR771181 | 9/12/23 | 2023.7 | 3III_B | Miami-Dade | Caribbean | 0.94 | Cluster 5 | 2023.31 | (2023.09,2023.54) |
| OR771185 | 9/16/23 | 2023.71 | 3III_B | Miami-Dade | Caribbean | 0.99 |  | 2023.63 | (2023.47,2023.7) |
| OR771182 | 9/20/23 | 2023.72 | 3III_B | Miami-Dade | Caribbean | 1 | Cluster 4 | 2023.27 | (2023.09,2023.45) |
| OR821936 | 9/21/23 | 2023.72 | 3III_B | Miami-Dade | Caribbean | 0.99 |  | 2023.63 | (2023.47,2023.7) |
| OR821940 | 9/19/23 | 2023.72 | 3III_B | Miami-Dade | Caribbean | 1 | Cluster 4 | 2023.27 | (2023.09,2023.45) |
| OR771188 | 9/26/23 | 2023.73 | 2II_F | Miami-Dade | South-America | 1 | Cluster 2 | 2023.48 | (2023.28,2023.62) |
| OR771189 | 9/26/23 | 2023.73 | 2II_F | Miami-Dade | Caribbean | 0.95 |  |  |  |
| OR821944 | 9/26/23 | 2023.73 | 2II_F | Miami-Dade | South-America | 1 | Cluster 2 | 2023.48 | (2023.28,2023.62) |
| OR771195 | 9/26/23 | 2023.73 | 3III_B | Miami-Dade | Caribbean | 1 | Cluster 4 | 2023.27 | (2023.09,2023.45) |
| OR821941 | 9/26/23 | 2023.73 | 3III_B | Miami-Dade | Caribbean | 1 | Cluster 4 | 2023.27 | (2023.09,2023.45) |
| OR821946 | 9/28/23 | 2023.74 | 2II_F | Miami-Dade | South-America | 1 | Cluster 2 | 2023.48 | (2023.28,2023.62) |
| OR771190 | 9/29/23 | 2023.74 | 3III_B | Miami-Dade | Caribbean | 1 | Cluster 4 | 2023.27 | (2023.09,2023.45) |
| OR821942 | 9/27/23 | 2023.74 | 3III_B | Miami-Dade | Caribbean | 1 | Cluster 4 | 2023.27 | (2023.09,2023.45) |
| OR821945 | 9/28/23 | 2023.74 | 3III_B | Miami-Dade | Caribbean | 1 | Cluster 4 | 2023.27 | (2023.09,2023.45) |
| OR771193 | 9/30/23 | 2023.75 | 2II_F | Miami-Dade | Caribbean | 0.95 |  |  |  |
| OR771191 | 9/30/23 | 2023.75 | 3III_B | Miami-Dade | Caribbean | 1 | Cluster 4 | 2023.27 | (2023.09,2023.45) |
| OR771199 | 10/3/23 | 2023.75 | 3III_B | Miami-Dade | Caribbean | 1 | Cluster 4 | 2023.27 | (2023.09,2023.45) |
| OR821948 | 9/30/23 | 2023.75 | 3III_B | Miami-Dade | Caribbean | 1 | Cluster 4 | 2023.27 | (2023.09,2023.45) |
| OR821950 | 10/2/23 | 2023.75 | 3III_B | Miami-Dade | Caribbean | 1 | Cluster 4 | 2023.27 | (2023.09,2023.45) |
| OR771192 | 10/4/23 | 2023.76 | 3III_B | Miami-Dade | Caribbean | 0.94 | Cluster 5 | 2023.31 | (2023.09,2023.54) |
| OR821949 | 10/6/23 | 2023.76 | 3III_B | Miami-Dade | Caribbean | 1 | Cluster 4 | 2023.27 | (2023.09,2023.45) |
| OR821951 | 10/7/23 | 2023.76 | 3III_B | Miami-Dade | Caribbean | 1 | Cluster 4 | 2023.27 | (2023.09,2023.45) |
| OR821952 | 10/5/23 | 2023.76 | 3III_B | Miami-Dade | Caribbean | 1 | Cluster 4 | 2023.27 | (2023.09,2023.45) |
| OR821954 | 10/10/23 | 2023.77 | 3III_B | Miami-Dade | Caribbean | 0.94 | Cluster 5 | 2023.31 | (2023.09,2023.54) |
| OR821955 | 10/9/23 | 2023.77 | 3III_B | Miami-Dade | Caribbean | 1 | Cluster 4 | 2023.27 | (2023.09,2023.45) |
| OR821958 | 10/10/23 | 2023.77 | 3III_B | Miami-Dade | Caribbean | 1 | Cluster 4 | 2023.27 | (2023.09,2023.45) |
| OR821962 | 10/11/23 | 2023.78 | 2II_F | Miami-Dade | Caribbean | 1 |  |  |  |
| OR821964 | 10/14/23 | 2023.78 | 3III_B | Miami-Dade | Caribbean | 1 | Cluster 4 | 2023.27 | (2023.09,2023.45) |
| OR977072 | 10/13/23 | 2023.78 | 3III_B | Miami-Dade | Caribbean | 1 | Cluster 4 | 2023.27 | (2023.09,2023.45) |
| OR977076 | 10/17/23 | 2023.79 | 3III_B | Miami-Dade | Caribbean | 1 | Cluster 4 | 2023.27 | (2023.09,2023.45) |
| OR977080 | 10/20/23 | 2023.8 | 2II_F | Miami-Dade | Caribbean | 1 |  |  |  |
| OR977077 | 10/22/23 | 2023.81 | 3III_B | Miami-Dade | Caribbean | 1 | Cluster 4 | 2023.27 | (2023.09,2023.45) |
| OR977081 | 10/25/23 | 2023.81 | 3III_B | Miami-Dade | Caribbean | 0.94 | Cluster 5 | 2023.31 | (2023.09,2023.54) |
| OR977085 | 10/28/23 | 2023.82 | 3III_B | Miami-Dade | Caribbean | 0.94 | Cluster 5 | 2023.31 | (2023.09,2023.54) |
| OR977086 | 11/1/23 | 2023.83 | 3III_B | Miami-Dade | Caribbean | 1 | Cluster 4 | 2023.27 | (2023.09,2023.45) |
| OR977088 | 11/7/23 | 2023.85 | 3III_B | Miami-Dade | Caribbean | 0.94 | Cluster 5 | 2023.31 | (2023.09,2023.54) |
| PP692455 | 11/12/23 | 2023.86 | 3III_B | Miami-Dade | Caribbean | 0.94 | Cluster 5 | 2023.31 | (2023.09,2023.54) |
| OR977091 | 11/15/23 | 2023.87 | 3III_B | Miami-Dade | Caribbean | 0.94 | Cluster 5 | 2023.31 | (2023.09,2023.54) |
| OR977092 | 11/15/23 | 2023.87 | 3III_B | Miami-Dade | Caribbean | 0.94 | Cluster 5 | 2023.31 | (2023.09,2023.54) |
| PP709309 | 11/20/23 | 2023.88 | 1V_D | Hardee | North-America | 0.98 |  |  |  |
| OR977097 | 11/20/23 | 2023.88 | 3III_B | Miami-Dade | Caribbean | 0.94 | Cluster 5 | 2023.31 | (2023.09,2023.54) |
| OR977099 | 11/19/23 | 2023.88 | 3III_B | Miami-Dade | Caribbean | 1 | Cluster 4 | 2023.27 | (2023.09,2023.45) |
| PP692465 | 12/12/23 | 2023.95 | 3III_B | Miami-Dade | Caribbean | 0.89 |  |  |  |
| PP708029 | 1/2/24 | 2024 | 3III_B | Miami-Dade | Caribbean | 1 |  |  |  |
| PP708030 | 1/8/24 | 2024.02 | 3III_B | Miami-Dade | Caribbean | 0.94 | Cluster 5 | 2023.31 | (2023.09,2023.54) |
| PP708038 | 1/20/24 | 2024.05 | 3III_B | Miami-Dade | Caribbean | 1 | Cluster 4 | 2023.27 | (2023.09,2023.45) |
| PQ340822 | 6/25/24 | 2024.48 | 3III_B | Miami-Dade | Caribbean | 1 |  |  |  |
| PQ527045 | 8/13/24 | 2024.61 | 3III_B | Hillsborough | Caribbean | 1 |  |  |  |
| PQ617129 | 8/26/24 | 2024.65 | 3III_B | Pasco | Caribbean | 1 |  |  |  |
| PQ617146 | 9/8/24 | 2024.69 | 3III_B | Miami-Dade | Caribbean | 1 |  |  |  |
| PQ617120 | 9/25/24 | 2024.73 | 3III_B | Palm-Beach | Caribbean | 0.71 |  |  |  |

**Table S5.**
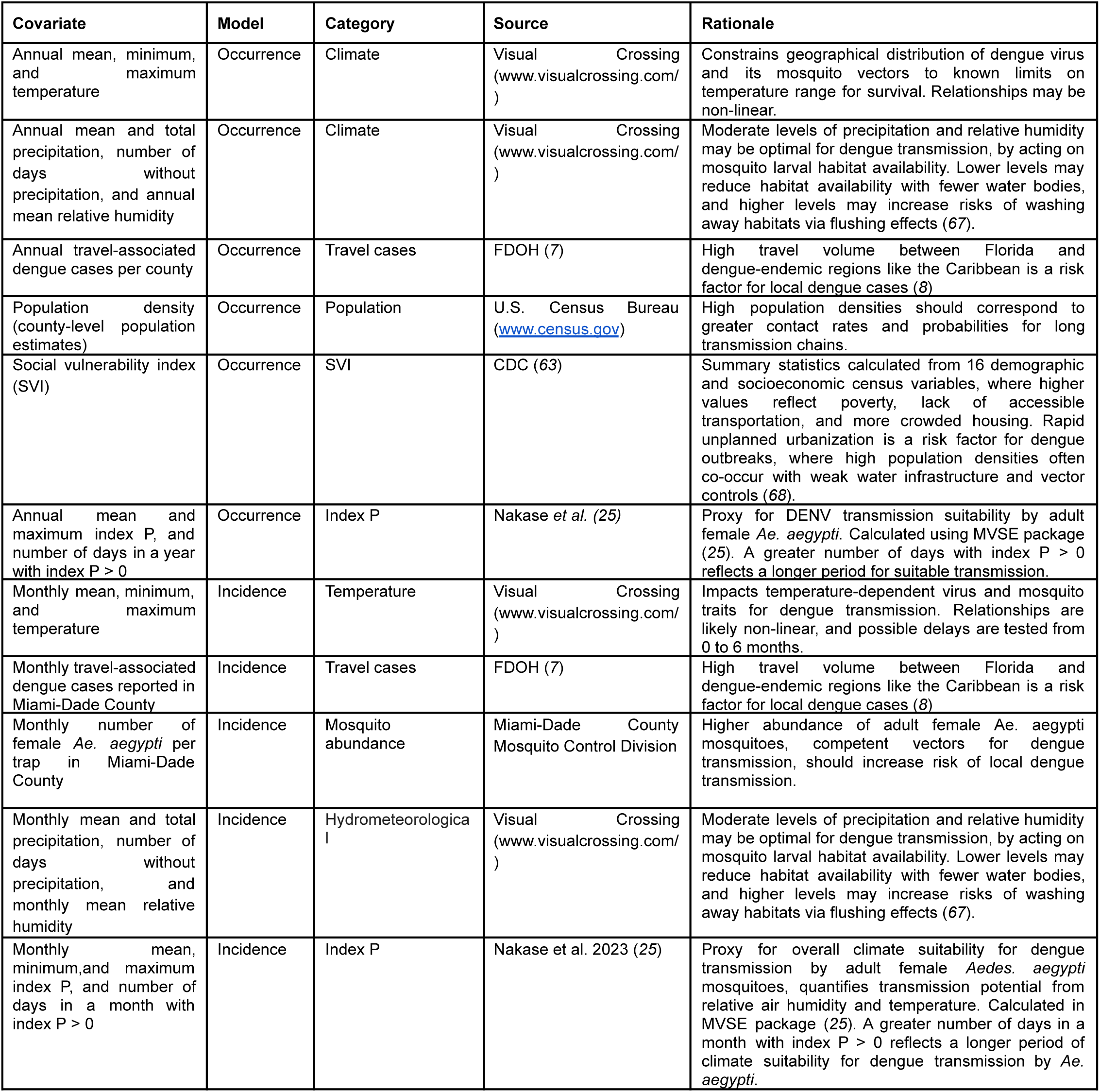
Covariates representing hypothesized drivers of local dengue infections in Florida.

**Table S6.**
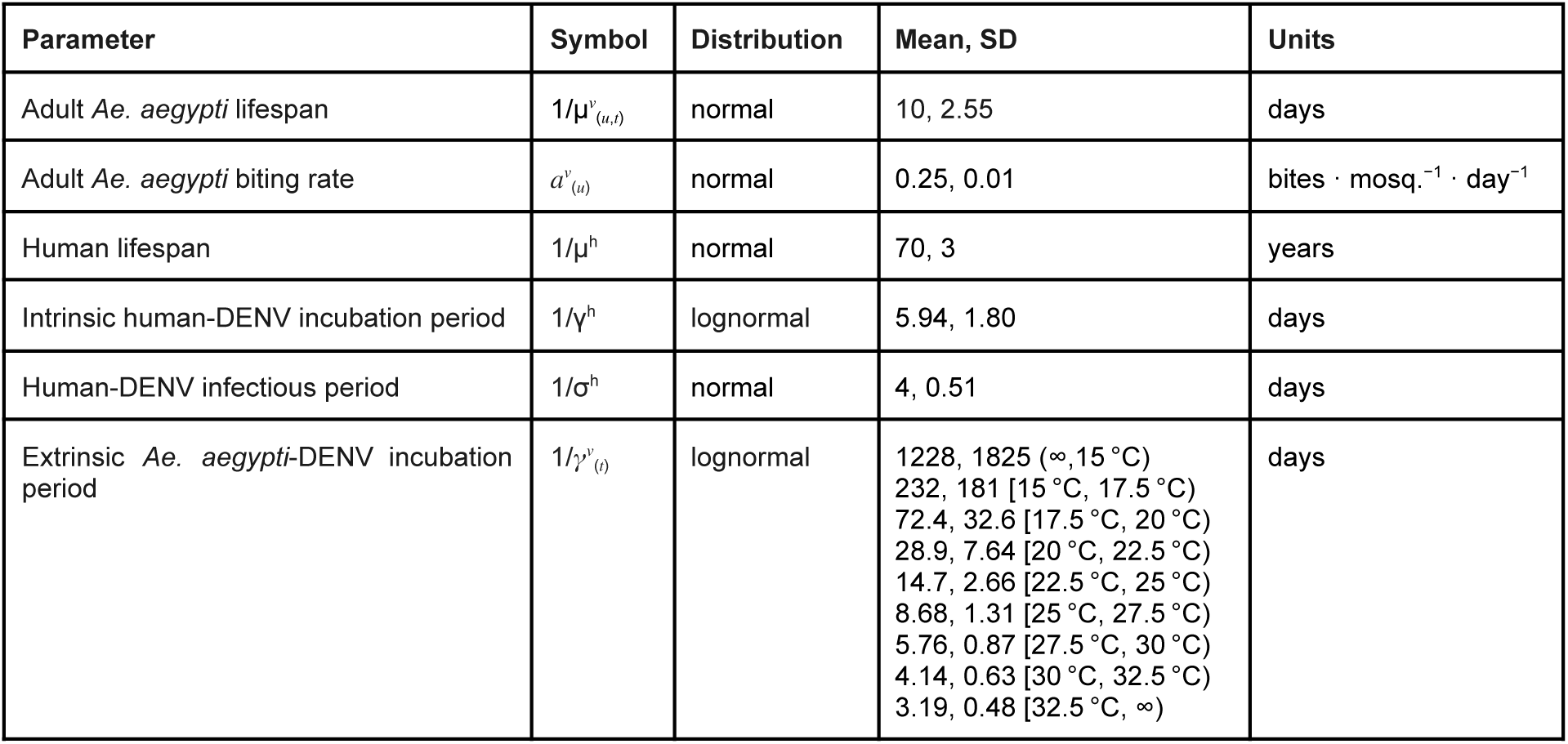
Default biological parameters used in estimating index P for DENV transmitted by *Ae. aegypti* mosquitoes from humidity (µ) and temperature (t) (from Nakase *et al.*)

| Parameter | Symbol | Distribution | Mean, SD | Units |
| --- | --- | --- | --- | --- |
| Adult *Ae. aegypti* lifespan | $1/\mu^v_{(\mu,t)}$ | normal | 10, 2.55 | days |
| Adult *Ae. aegypti* biting rate | $a^v_{(\mu)}$ | normal | 0.25, 0.01 | bites · mosq.<sup>-1</sup> · day<sup>-1</sup> |
| Human lifespan | $1/\mu^h$ | normal | 70, 3 | years |
| Intrinsic human-DENV incubation period | $1/\gamma^h$ | lognormal | 5.94, 1.80 | days |
| Human-DENV infectious period | $1/\sigma^h$ | normal | 4, 0.51 | days |
| Extrinsic *Ae. aegypti*-DENV incubation period | $1/\gamma^v_{(t)}$ | lognormal | 1228, 1825 (∞, 15 °C)<br>232, 181 [15 °C, 17.5 °C)<br>72.4, 32.6 [17.5 °C, 20 °C)<br>28.9, 7.64 [20 °C, 22.5 °C)<br>14.7, 2.66 [22.5 °C, 25 °C)<br>8.68, 1.31 [25 °C, 27.5 °C)<br>5.76, 0.87 [27.5 °C, 30 °C)<br>4.14, 0.63 [30 °C, 32.5 °C)<br>3.19, 0.48 [32.5 °C, ∞) | days |

**Table S7.**
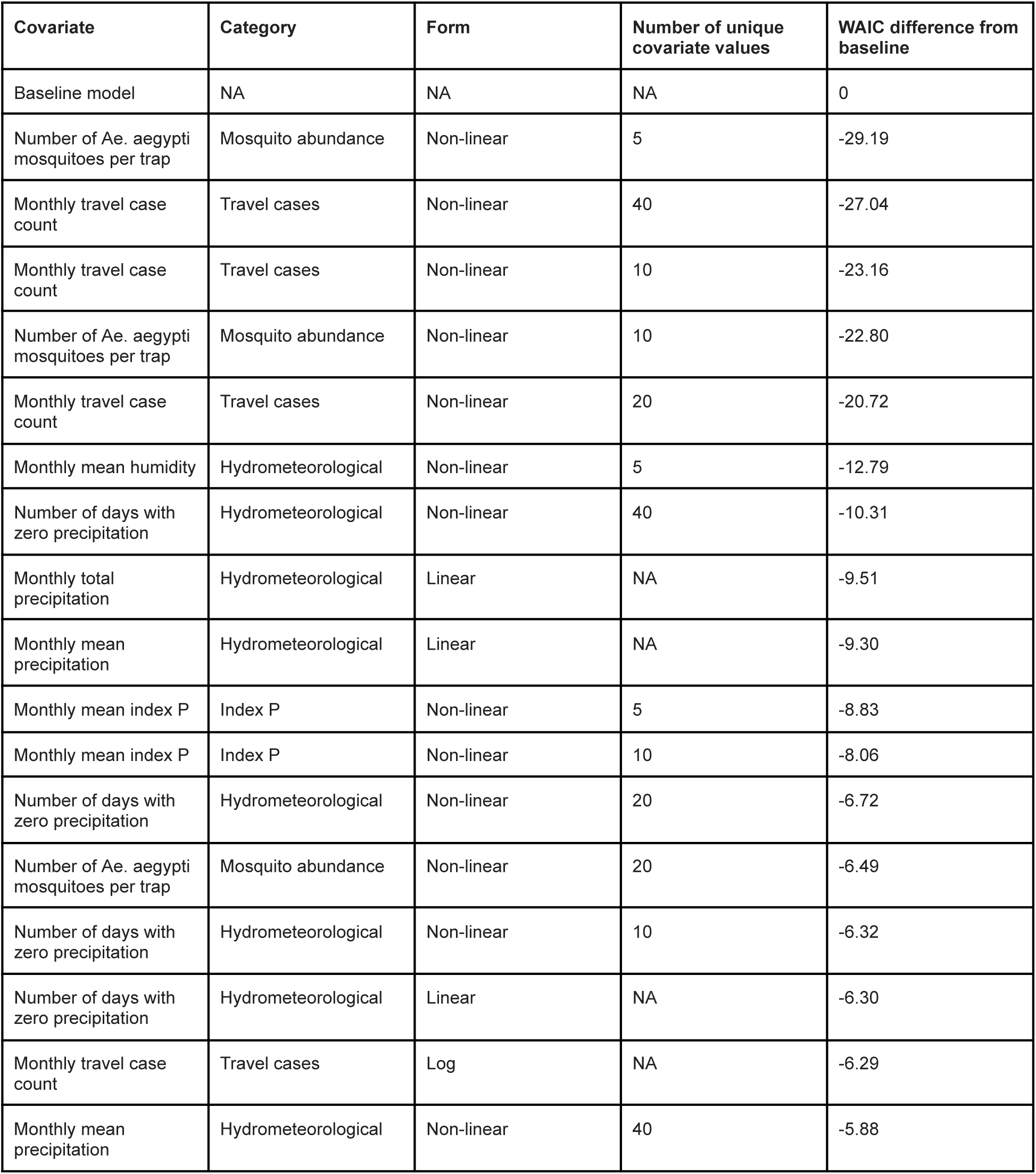
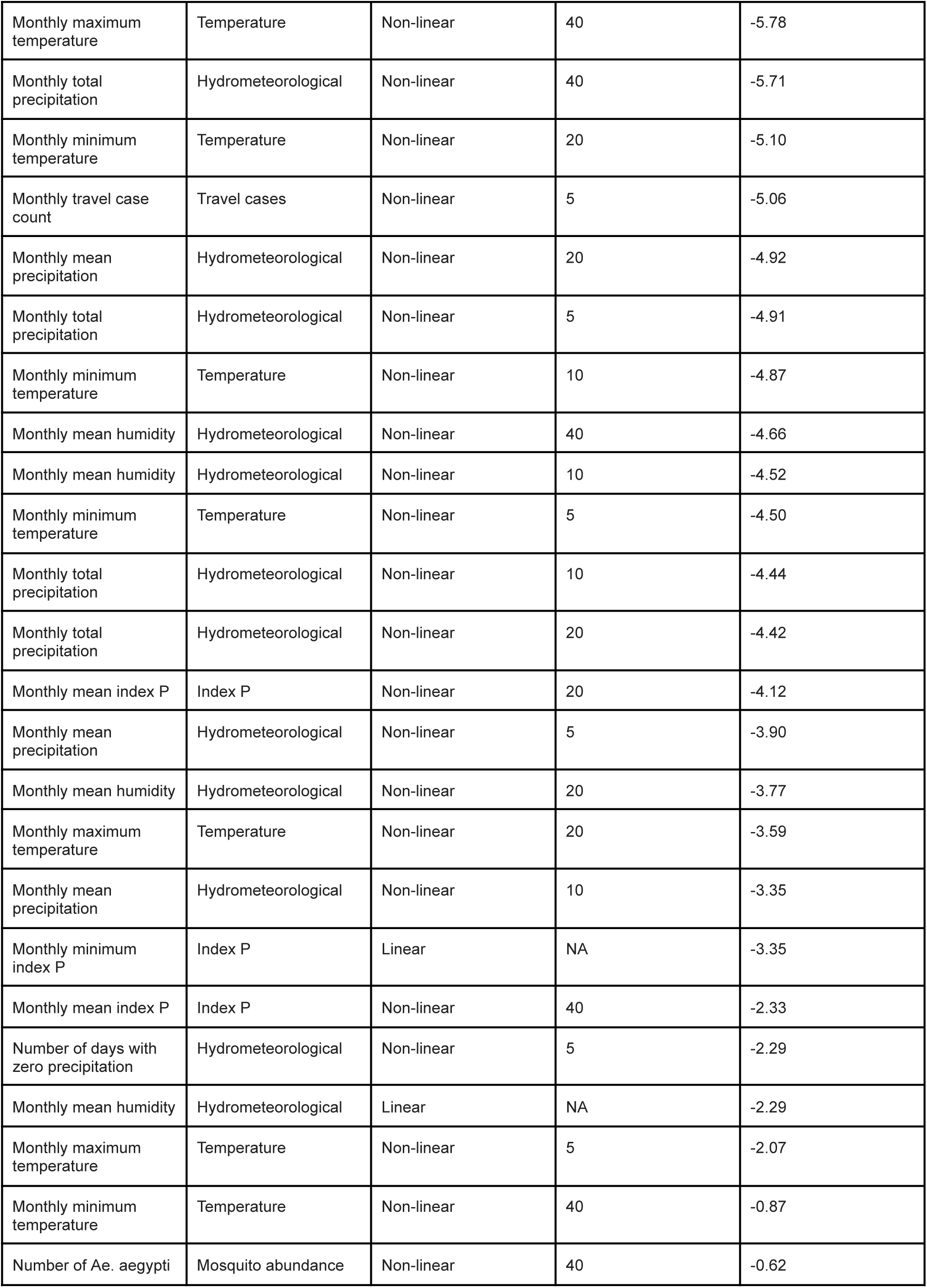
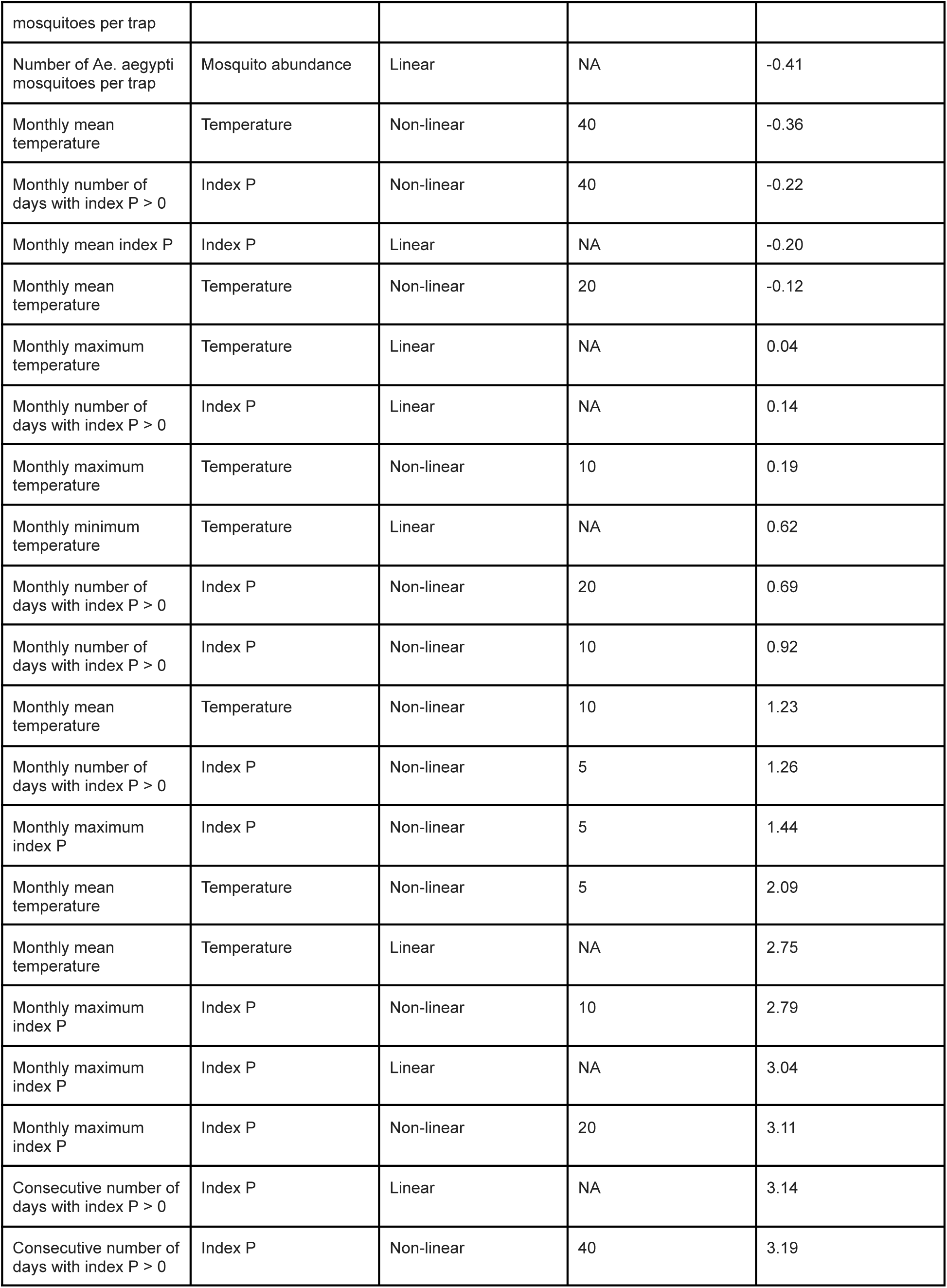
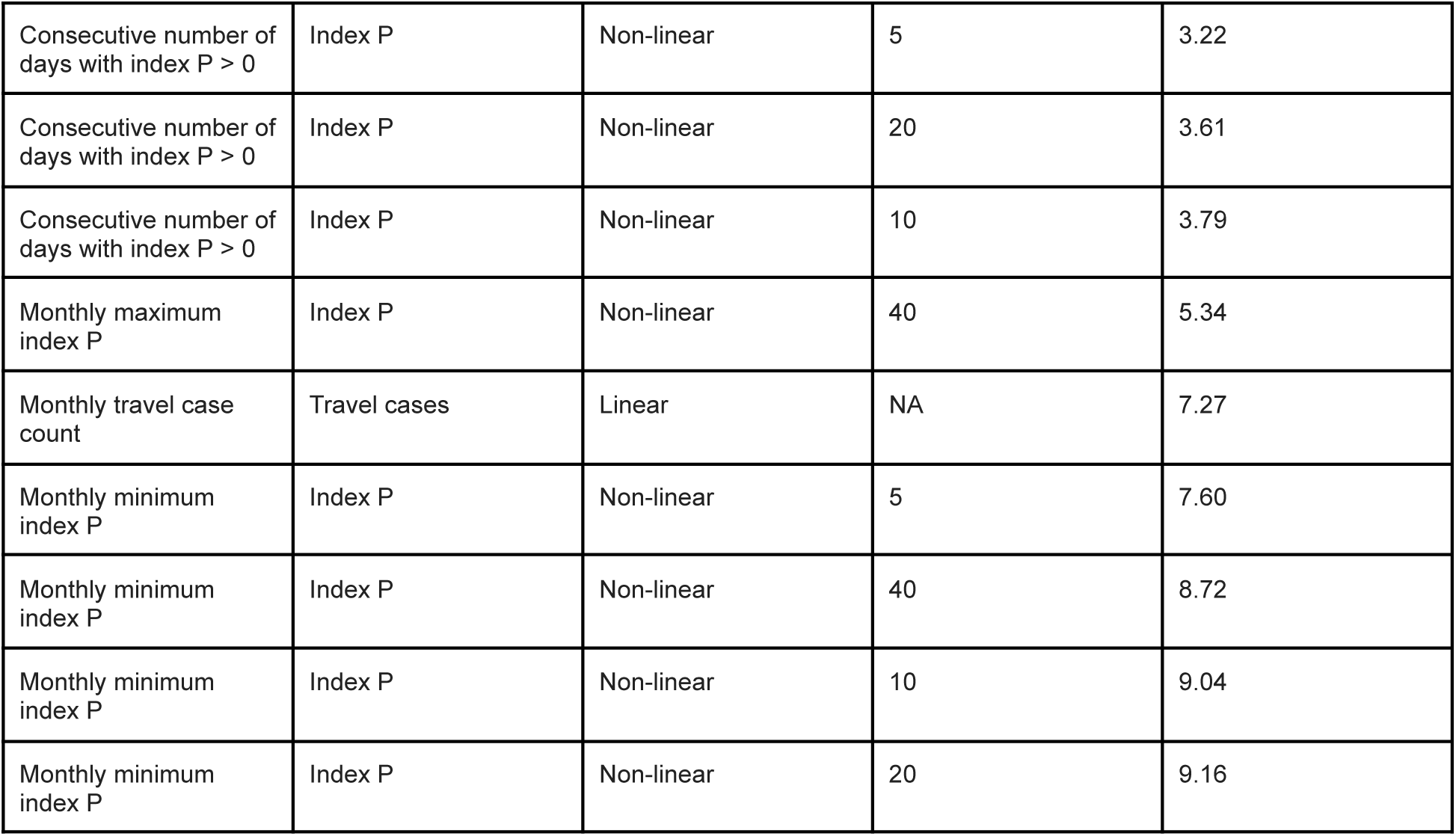
Summary metrics for baseline and univariate models of monthly local dengue case incidence in Miami-Dade County from 2009 to 2024, the latter fitted as a baseline model with the inclusion of a single covariate. Models are ordered by ascending WAIC values, where larger values indicate a worse fit to the observed data. Covariates are categorized by type and functional form. Non-linear effects were modeled as a second-order random walk over a varying number of unique covariate values (5, 10, 20, and 40), where a greater number allowed for more flexibility.

## Notes

### Competing Interest Statement

The authors have declared no competing interest.

### Summary of Updates

Author name corrected; no other changes.

